# Assessing and refining a metabolite-based distress score for use in different populations within three US cohorts

**DOI:** 10.64898/2026.09.10.26362734

**Authors:** Tianyi Huang, Yiwen Zhu, Raji Balasubramanian, Andrea L. Roberts, Clary B. Clish, Julian Avila-Pacheco, Jerome I. Rotter, Xiuqing Guo, Jie Yao, Yii-Der Ida Chen, Stephen S. Rich, Kent D. Taylor, Alexis C. Wood, W. Craig Johnson, Katherine H. Shutta, Kathryn M. Rexrode, Aladdin H. Shadyab, Su Yon Jung, JoAnn E. Manson, Susan E. Hankinson, Laura D. Kubzansky

**Affiliations:** Laboratory of Epidemiology and Population Sciences, Intramural Research Program, National Institute on Aging, Baltimore, MD, USA; Department of Epidemiology, Harvard T.H. Chan School of Public Health, Boston, MA, USA; Department of Biostatistics and Epidemiology, School of Public Health and Health Sciences, University of Massachusetts Amherst, Amherst, MA, USA; Department of Environmental Health, Harvard T.H. Chan School of Public Health, Boston, MA, USA; Broad Institute of Massachusetts Institute of Technology and Harvard, Cambridge, MA, USA; The Institute for Translational Genomics and Population Sciences, Department of Pediatrics, The Lundquist Institute for Biomedical Innovation at Harbor-UCLA Medical Center, Torrance, CA, USA; Department of Genome Sciences, University of Virginia, Charlottesville, VA, USA; USDA/ARS Children’s Nutrition Research Center, Baylor College of Medicine, Houston, TX, USA; Department of Biostatistics, University of Washington School of Public Health, Seattle, WA, USA; Department of Biostatistics, Harvard T.H. Chan School of Public Health, Boston, MA, USA; Division of Women’s Health, Department of Medicine, Mass General Brigham, Harvard Medical School, Boston, MA, USA; Herbert Wertheim School of Public Health and Human Longevity Science and Division of Geriatrics, Gerontology, and Palliative Care, Department of Medicine, University of California, San Diego, La Jolla, CA, USA; Jonsson Comprehensive Cancer Center, Translational Sciences Section, School of Nursing, University of California, Los Angeles, CA, USA; Department of Epidemiology, Fielding School of Public Health, University of California, Los Angeles, Los Angeles, CA, USA; Division of Preventive Medicine, Brigham and Women’s Hospital, Harvard Medical School, Boston, MA, USA; Department of Social and Behavioral Sciences, Harvard T.H. Chan School of Public Health, Boston, MA, USA

## Abstract

**Introduction:** Psychological distress is associated with metabolic alterations that contribute to adverse cardiometabolic outcomes across diverse populations. We previously developed a metabolite-based distress score (MDS) in predominantly White women. Because men and Black individuals suffer high rates of cardiometabolic disease, we evaluated whether the MDS is similarly associated with depression in these populations, and derived a multi-ethnic MDS (MDS-ME) among Black individuals with replications in independent samples.

**Methods:** Data were from three U.S. cohorts, the Multi-Ethnic Study of Atherosclerosis (MESA), Women’s Health Initiative (WHI), and Nurses’ Health Study (NHS), evaluated in 4 stages. Stages 1-3 included 2,477 White and 1,625 Black MESA participants with depression status data and plasma metabolomics assessed at baseline. Stage 1: We calculated the MDS and examined its association with depression status. Stage 2: We conducted an agnostic analysis among a 70% random subset of Black participants to identify additional relevant metabolites. Stage 3: We derived an MDS-ME in Black participants. Stage 4: We evaluated the MDS-ME using independent samples from all three cohorts.

**Results:** The previously validated 20-metabolite MDS showed stronger associations with prevalent depression in White (OR: 1.93, 95% CI: 1.71, 2.19) versus Black (OR: 1.13, 95% CI: 0.97, 1.33) MESA participants. Associations were generally similar in men and women. The MDS-ME comprised 33 metabolites, and relative to the MDS was more strongly associated with depression in Black participants (OR: 1.45; 95% CI: 1.04, 2.03), with associations also evident in White participants (OR: 1.81; 95% CI: 1.60, 2.04). The MDS-ME was similarly associated with depression in Black women in WHI and NHS.

**Conclusion:** Results suggest that incorporating metabolites identified in Black participants enhances the utility and generalizability of the MDS across more diverse populations.

## Introduction

Multiple forms of psychological distress, such as depression, are increasingly recognized as key risk factors for cardiometabolic disease and as contributors to health disparities (Djuric et al., 2008; Talala et al., 2011; Turner, 2013; Vaccarino et al., 2025). The ways in which psychological distress is experienced, expressed, and biologically embedded may vary across population groups. Cultural norms, social contexts, and gender roles can influence how distress is perceived and reported, as well as how distress may engage downstream biologic pathways (Assari, 2017; Lincoln et al., 2011; Pamplin and Bates, 2021). Such norm- and context-related variation may differ across racial groups and between men and women, potentially contributing to heterogeneity in the biological correlates of psychological distress. However, the pathways linking psychological distress to cardiometabolic risk, including distress-associated metabolic profiles that may differ by race and sex, remain poorly understood.

Metabolic alterations are important biological correlates of psychological distress and may provide insight into pathways through which distress promotes cardiometabolic disease risk. Metabolomic studies have shown that perturbations in amino acid, lipid, and energy metabolism are associated with depressive symptoms and chronic stress, suggesting that metabolic pathways may serve as key mediators leading to adverse physical health outcomes (Bot et al., 2020; Caspani et al., 2021; Huang et al., 2021; Miao et al., 2023; Shutta et al., 2021; Zhu et al., 2022). In recent work in predominantly White women, we identified and validated a metabolite-based distress score (MDS) consisting of 20 plasma metabolites that showed robust differences across levels of psychological distress captured by a composite measure of depression and anxiety (Balasubramanian et al., 2023; Shutta et al., 2021). Further, the MDS was associated with increased risk of incident diabetes and cardiovascular disease among predominantly White men and women, independent of conventional cardiometabolic risk factors (Balasubramanian et al., 2023; Huang et al., 2024). However, this MDS has not yet been evaluated in more diverse populations, particularly among Black individuals or men, populations that suffer high rates of cardiometabolic disease.

Interestingly, a growing body of evidence highlighted considerable differences in plasma metabolomic profiles between Black and White individuals, which have been partly attributed to behavioral and environmental factors (Butler et al., 2023; Hu et al., 2022; McGee et al., 2024). Notable metabolomic differences were also observed between men and women, with patterns that varied dynamically with age (Bell et al., 2021; Costanzo et al., 2022). It is possible that the MDS previously developed in primarily White, post-menopausal women may not fully reflect distress-related metabolic profiles in Black individuals and men, which may be partly attributable to differences in socioenvironmental exposures, lifestyles, stress physiology, and/or genetic or epigenetic regulation.

Therefore, the objectives of the current study are (1) to evaluate the association of our previously developed MDS with depression in White and Black men and women and (2) as appropriate, to evaluate if additional distress-related metabolites might better capture a metabolite profile of distress in these groups by deriving a multi-ethnic MDS (MDS-ME). To address these objectives, we conducted this work across multiple U.S. cohorts, including the Multi-Ethnic Study of Atherosclerosis (MESA), the Women’s Health Initiative (WHI) and the Nurses’ Health Study (NHS). The study was conducted in four stages to evaluate, identify, revise, and replicate the metabolomic signature of distress in Black participants, and secondarily, to assess potential sex differences (**Figure 1**). This multi-cohort, multi-stage approach allows us to build upon our prior work, assess the consistency of observed associations, identify cohort-specific differences, and strengthen the robustness and reproducibility of the results. Understanding racial and sex differences in the metabolic manifestations of distress is critical for advancing mechanistic insight into health disparities.

**Figure 1.**
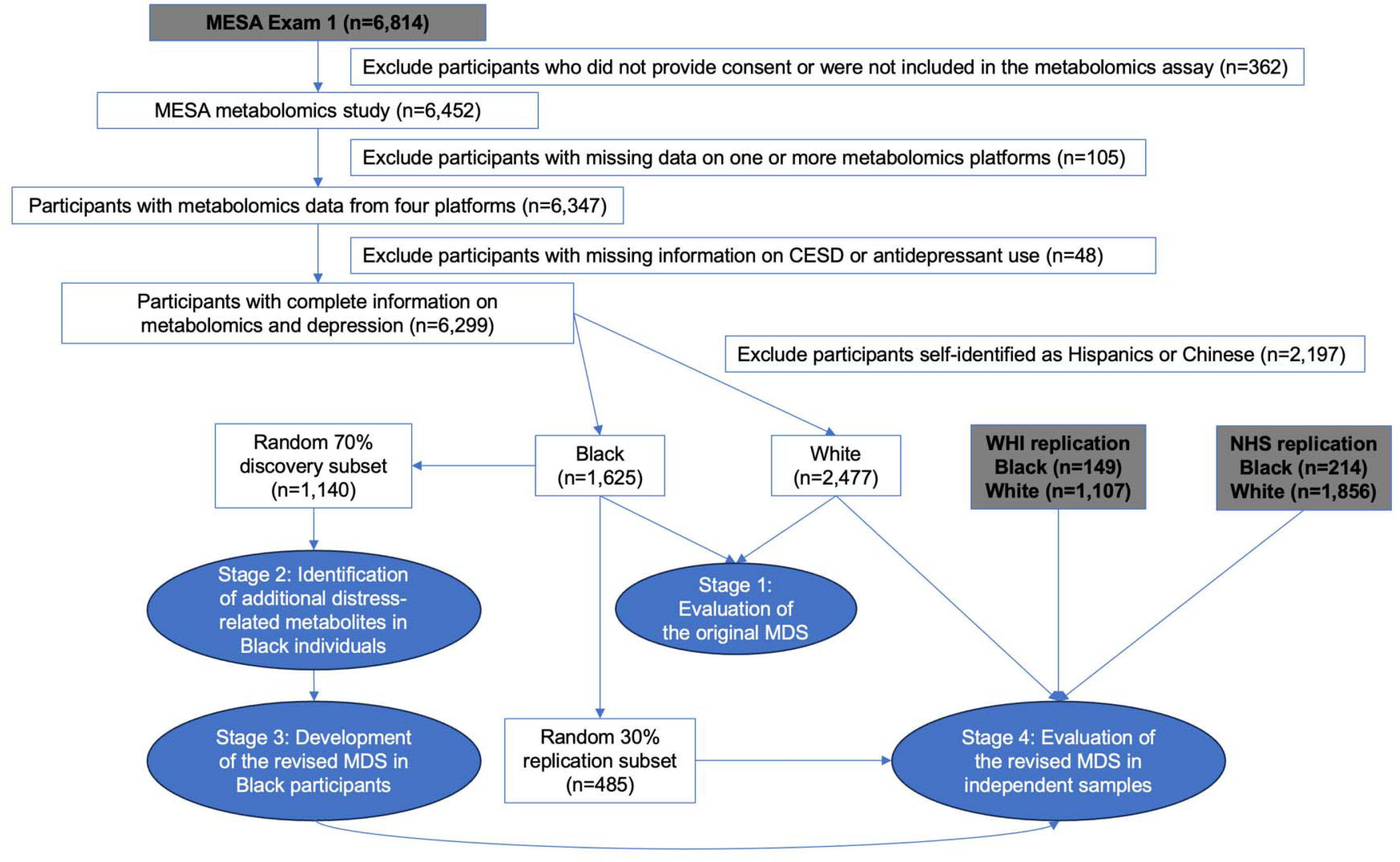
Flow chart for the analytic samples. Note that our analysis was restricted to Black and White participants in the Multi-Ethnic Study of Atherosclerosis (MESA), as they were the ancestry groups for which replication was possible in the Women’s Health Initiative (WHI and the Nurses’ Health Study (NHS). Participants from other ethnic groups in MESA (Hispanics and Chinese) were not included in this study.

## Methods

### Study population

Data were from three US cohorts (MESA, WHI, and NHS). MESA is a prospective observational cohort that recruited 6,814 participants free of cardiovascular disease from six US communities in 2000-2004 (baseline age: 45-84 years) (Bild et al., 2002). The WHI consists of 161,808 postmenopausal women who were recruited from 40 clinical centers across the US in 1993-1998 (baseline age: 50-79 years) (1998). The NHS is an ongoing, longitudinal cohort of 121,700 female registered US nurses who completed biennial follow-up surveys since enrollment in 1976 (baseline age: 30-55 years) (Colditz and Hankinson, 2005). In all cohorts, race was self-reported by participants.

In our four-stage study (**Figure 1**), Stages 1-3 use data from 2,477 White and 1,625 Black MESA participants with complete data on plasma metabolomics and depression-related measures. The analysis was further restricted to a 70% training subset of Black participants in Stages 2-3 to derive the MDS-ME. In Stage 4, to provide additional independent assessment of the MDS-ME in MESA, we utilized the remaining withheld 30% subset of MESA Black participants, as well as additional Black and White women from WHI and NHS for whom at least a subset of the newly identified metabolites in the MDS-ME were available. In WHI, our analytic sample included 149 Black women and 1,107 White women who participated in the Observational Study (WHI-OS) and were included in a previous nested case-control study of plasma metabolomics and coronary heart disease (Paynter et al., 2018). The NHS analysis included 214 Black women and 1856 White women who were included in a prior analysis of racial differences in metabolomic profiles (McGee et al., 2024). Of note, while the original 20-metabolite MDS was developed in samples of primarily White women from WHI and NHS, the replication samples included in the current study were independent, with no overlap with our prior analyses.

### Depression assessment

In <u>MESA</u>, following previous work in this area, depression status at baseline was defined by the presence of one or both of the following: (1) current antidepressant use or (2) a score ≥16 on the 20-item Center for Epidemiologic Studies Depression Scale (CES-D), an established cutoff for probable depression (Radloff, 1977). In <u>WHI</u> and <u>NHS</u>, depression status was ascertained according to use of antidepressants or obtaining a score above the specified cut point on a validated depressive symptom measure, with a third element used in each dataset to capture self-reported depression history. Specifically, in <u>WHI</u>, depression was defined as meeting any of the following: (1) current antidepressant use, (2) CES-D score ≥16, or (3) self-reported history of depressed mood. In <u>NHS</u>, using depression measures obtained on surveys closest to blood collection (range 2-6 years after blood collection), we defined depression as: (1) current antidepressant use, (2) scoring ≤52 on the 5-item Mental Health Index (MHI-5), an established cut point previously used with this measure,(Berwick et al., 1991) or (3) self-reported history of receiving a clinician’s diagnosis of depression.

### Metabolomic measurement

Plasma metabolomics profiling was performed at the Broad Institute of the Massachusetts Institute of Technology and Harvard using liquid chromatography tandem mass spectrometry (LC-MS) platforms for all three samples. In <u>MESA</u>, data were generated from four platforms: (1) the C8-positive platform primarily detects moderately hydrophobic metabolites including lipids and sterol derivatives, (2) the C18-negative platform is optimized for compounds of intermediate polarity, such as fatty acids, bile acids and complex lipids, (3) the HILIC-positive platform measures polar, water-soluble metabolites, including amino acids, carnitines, and biogenic amines, and (4) the HILIC-negative platform captures polar and acidic metabolites such as organic acids and carbohydrates. Together, these platforms capture a broad range of identified metabolites (n=764 metabolites). Details on profiling methods have been described previously (Clish, 2015; Paynter et al., 2018).

In <u>WHI</u>, metabolomics data were available for the same four platforms; however, because these data were generated several years prior to the MESA profiling, they featured a smaller number of annotated metabolites identified at the time (n=472 metabolites) (Hu et al., 2022; Paynter et al., 2018). In <u>NHS</u>, metabolomics data were available from only two platforms (HILIC-positive and C8-positive) and also had limited coverage (n=334) (McGee et al., 2024).

### Covariates

We examined two sets of covariates to provide estimates adjusted for key confounders (Model 1) and for a wider range of medical and biobehavioral factors (Model 2). In MESA, covariates were assessed at baseline, concurrent with blood collection. Model 1 included age (years), sex, study site, education (high school or less, some college, college graduate, graduate school), employment status (homemaker, employed, unemployed, retired), and household income (<$20,000, $20,000-$49,999, $50,000-$99,999, ≥$100,000). Model 2 further adjusted for BMI (continuous), smoking status (current, former, never), physical activity (MET hrs/week, continuous), diet quality scores (continuous), systolic blood pressure (continuous), HDL cholesterol (continuous), total cholesterol (continuous), diabetes status (yes, no), and use of antihypertensive and lipid-lowering medications (yes, no). We attempted to adjust for comparable sets of covariates in WHI and NHS; however, a small number of covariates could not be included because of data limitations. Details on covariate assessment are provided in **Supplemental Table 1**.

### Statistical analysis

#### Stage 1: Evaluation of the original MDS

Using data from MESA White and Black participants, we calculated the MDS based on 20 metabolites previously identified (**Supplemental Table 2**).^14^ Each metabolite was log-transformed and z-scored separately among White and Black participants before being weighted by its corresponding coefficient to generate the 20-metabolite MDS, which was then z-scored within each racial group. Using logistic regression, depression status was regressed on the standardized MDS, such that the odds ratio (OR) represented the odds of probable depression per 1-SD increase in the MDS. All analyses were conducted separately in White and Black participants, and within each racial group we further stratified by sex to evaluate potential differences between men and women. Statistical interactions by sex were assessed using a likelihood ratio test comparing models with and without the cross-product term. Two multivariable-adjusted models were considered in these analyses as described above.

#### Stage 2: Identification of additional distress-related metabolites in Black individuals

We randomly divided the MESA Black participants into two subsets (70% for discovery and 30% for replication). We conducted an agnostic analysis assessing associations between depression and 764 metabolites in the 70% discovery subset (n=1,140) to identify additional metabolites potentially associated with depression among Black participants beyond the 20 metabolites included in the original MDS. Following a similar strategy used for developing the original MDS, we used logistic regression to estimate the odds ratio (OR) and 95% CI of probable depression for each metabolite (standardized to z-scores), applying the same two multivariable-adjusted models described above. Because our primary objective was to identify informative metabolites that may enhance the original MDS in Black participants rather than to make definitive causal inferences, we used a liberal inclusion threshold (p<0.05) to select candidate metabolites, prioritizing sensitivity over strict control of false positives.

#### Stage 3: Development of the MDS-ME in MESA Black participants

Metabolites nominally associated with depression in the previous step (p<0.05) were considered as candidates for the MDS-ME. Further, to build the MDS-ME based on our prior work, all 20 metabolites included in the original MDS were also included as candidates regardless of whether their associations reached the nominal significance level among Black participants. To select metabolites, candidate metabolite z-scores were entered into a LASSO logistic regression model as explanatory variables, with depression status as the response variable (Friedman et al., 2010; Tibshirani, 1996). Model tuning was performed using 10-fold cross-validation to determine the optimal penalty parameter that minimized the cross-validated deviance. Metabolites with nonzero coefficients in the optimal model were retained. The MDS-ME was calculated as the weighted sum of these selected metabolites using their LASSO-derived coefficients.

#### Stage 4: Evaluation of the MDS-ME with depression in three independent samples

To evaluate reproducibility and generalizability, we evaluated the association of the MDS-ME with depression in the independent 30% replication set of MESA Black participants (n=485). We also evaluated the performance of the MDS-ME among MESA White participants (n=2,477) and assessed potential sex differences in White and Black men and women. To assess the generalizability of the MDS-ME in other cohorts, we evaluated the MDS-ME in independent samples from the WHI (149 Black and 1,107 White women) and NHS (214 Black and 1856 White women). Of note, only a subset of metabolites identified for the MDS-ME in MESA were available in WHI and NHS, with slightly different subsets available in each cohort. We recreated a reduced MDS-ME in MESA by including only metabolite subsets that were available in WHI and NHS, and compared them with the full MDS-ME in MESA. We adjusted for comparable sets of covariates in WHI and NHS as possible; however, a small number of covariates could not be included because of data limitations (**Supplemental Table 1**). We implemented random-effects meta-analysis of the fully-adjusted estimates of the associations of MDS-ME with depression among Black participants, with inference based on the Hartung-Knapp-Sidik-Jonkman method to assess potential heterogeneity across cohorts (IntHout et al., 2014). All analyses were conducted in RStudio (version 4.3.5).

## Results

### Sample characteristics

In <u>MESA</u>, the mean age (SD) was 62.6 (10.2) years in 2,477 White participants and 62.2 (10.1) years in 1,625 Black participants (**Table 1**). Despite slightly higher depressive symptoms (both continuous and binary CESD measures) in Black compared with White participants, antidepressant use was more common in White participants. As a result, probable depression was more prevalent in White (19.9%) than in Black participants (14.3%). Compared with White participants, Black participants tended to report lower educational attainment and household income, unemployment or retirement, current smoking, higher physical activity and poorer diet quality. On average, Black versus White participants exhibited higher systolic blood pressure and greater use of antihypertensive medications, lower total cholesterol, and less use of lipid-lowering medications. BMI was higher and diabetes was more prevalent among Black participants. Similar differences were observed between White and Black women in <u>WHI</u> and <u>NHS</u> (**Supplemental Table 3**).

**Table 1.**
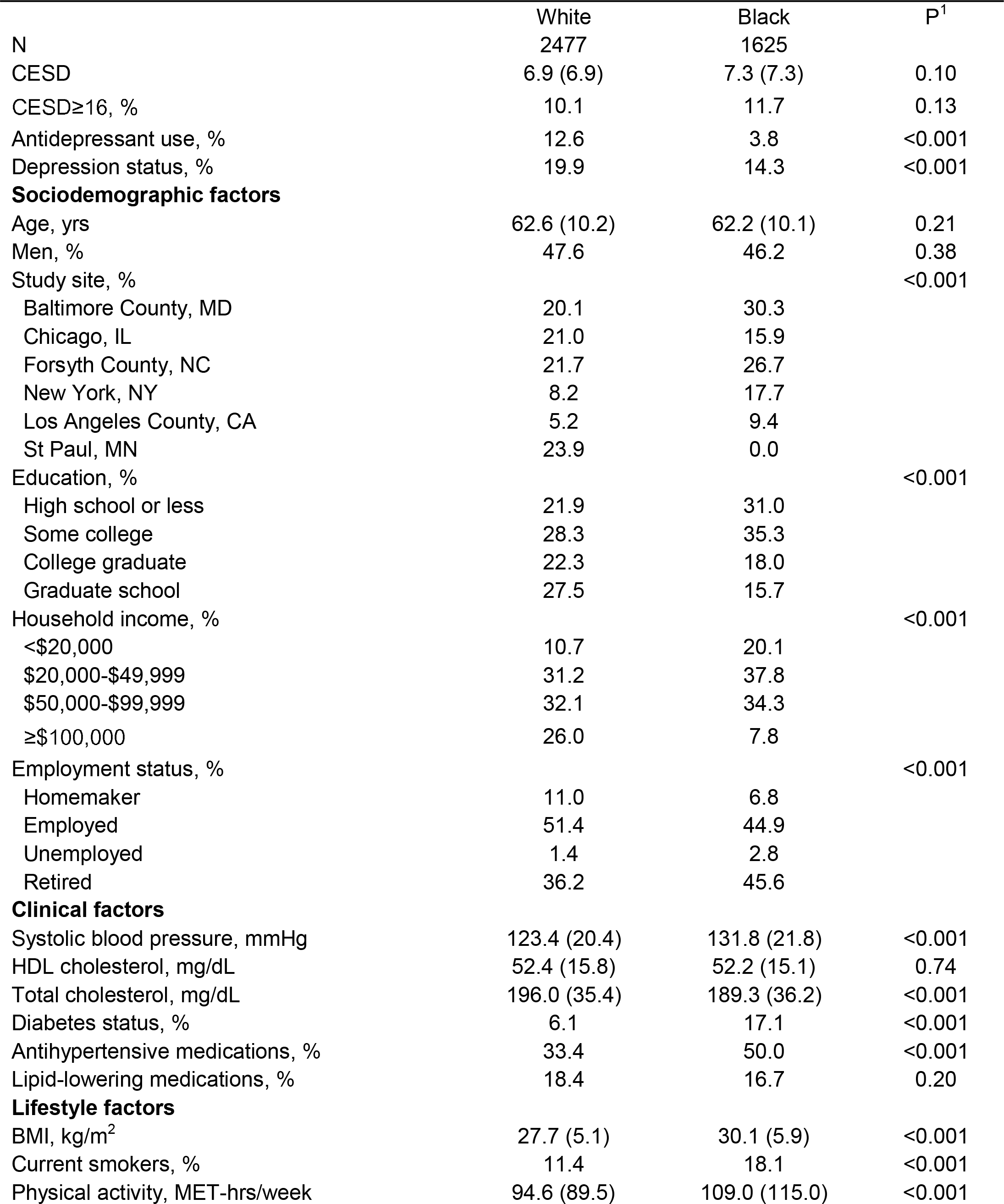
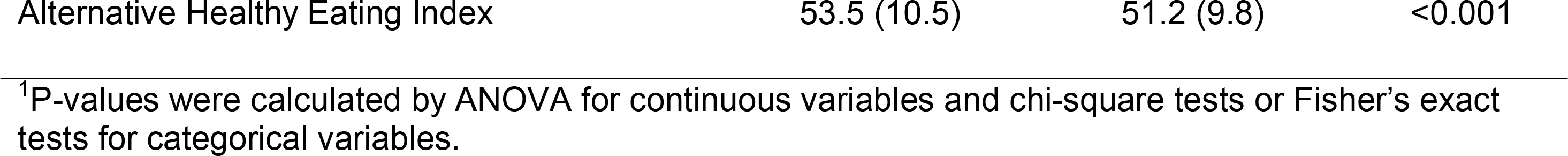
Sample characteristics in White and Black participants from MESA.

### Stage 1: Evaluation of the original MDS

The 20-metabolite MDS, previously developed from independent samples of primarily White women, was strongly associated with depression status in MESA White participants (**Table 2**). After adjusting for age, sex, study site, education, household income and employment status, each 1-SD increment in the MDS was associated with 86% higher odds of depression (95% CI: 1.67, 2.09). The association became somewhat stronger with further adjustment of clinical and lifestyle factors (OR: 2.00; 95% CI: 1.77, 2.25). When examining associations stratified by sex, no appreciable differences were observed between White women (n=1,298) and men (n=1,179) in either model (p-interaction>0.88; **Table 2**). In contrast, the association between the original MDS and depression status was substantially weaker among Black participants (comparable OR: 1.16; 95% CI: 1.00, 1.35) after adjusting for sociodemographic factors and remained essentially the same after additionally adjusting for clinical and lifestyle factors. When considering associations in Black men (n=750) and women (n=875) separately, the association appeared stronger in men (OR: 1.35; 95% CI: 1.03, 1.78) but was null in women (OR: 1.06; 95% CI: 0.86, 1.30; p-interaction=0.08).

**Table 2.**
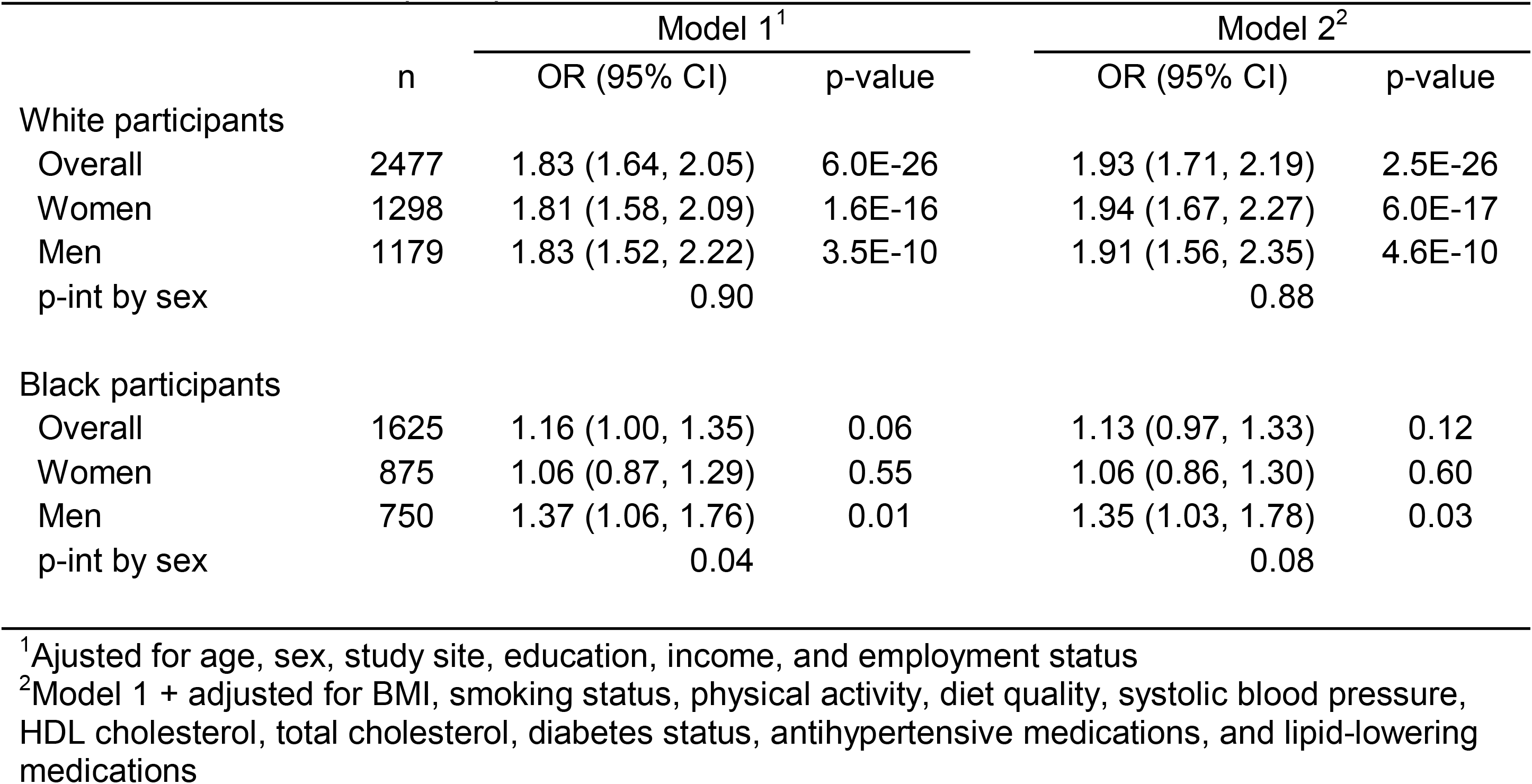
Associations between the original 20-metabolite MDS (per 1-SD increment) and depression status in White and Black participants from MESA.

| Status in White and Black participants from MESA |  |  |  |  |  |
| --- | --- | --- | --- | --- | --- |
|  |  | Model 1<sup>1</sup> |  | Model 2<sup>2</sup> |  |
|  | n | OR (95% CI) | p-value | OR (95% CI) | p-value |
| White participants |  |  |  |  |  |
| Overall | 2477 | 1.83 (1.64, 2.05) | 6.0E-26 | 1.93 (1.71, 2.19) | 2.5E-26 |
| Women | 1298 | 1.81 (1.58, 2.09) | 1.6E-16 | 1.94 (1.67, 2.27) | 6.0E-17 |
| Men | 1179 | 1.83 (1.52, 2.22) | 3.5E-10 | 1.91 (1.56, 2.35) | 4.6E-10 |
| p-int by sex |  | 0.90 |  | 0.88 |  |
| Black participants |  |  |  |  |  |
| Overall | 1625 | 1.16 (1.00, 1.35) | 0.06 | 1.13 (0.97, 1.33) | 0.12 |
| Women | 875 | 1.06 (0.87, 1.29) | 0.55 | 1.06 (0.86, 1.30) | 0.60 |
| Men | 750 | 1.37 (1.06, 1.76) | 0.01 | 1.35 (1.03, 1.78) | 0.03 |
| p-int by sex |  | 0.04 |  | 0.08 |  |

<sup>1</sup>Adjusted for age, sex, study site, education, income, and employment status
<sup>2</sup>Model 1 + adjusted for BMI, smoking status, physical activity, diet quality, systolic blood pressure, HDL cholesterol, total cholesterol, diabetes status, antihypertensive medications, and lipid-lowering medications

### Stage 2: Identification of additional distress-related metabolites in Black individuals

Given the weaker association between the original MDS and depression among Black participants and especially in Black women, we performed an agnostic analysis by examining the associations of individual 764 metabolites with depression in a subset of MESA Black participants (n=1,140; **Supplemental Table 4**). In Model 1, 57 metabolites were nominally associated with depression, while in Model 2 (fully-adjusted) 46 metabolites were nominally associated of which 41 were identified in Model 1. Based on Model 2 (**Figure 2**), the three metabolites most strongly associated with lower odds of depression were serotonin (OR: 0.64, 95% CI: 0.53, 0.77), hydroxytryptophan (OR: 0.67, 95% CI: 0.57, 0.80), and adenosine (OR: 0.73, 95% CI: 0.62, 0.87). Other metabolites inversely associated with depression were primarily various carnitine species. Metabolites positively associated with depression mainly included PCs, LPCs, LPEs, long-chain saturated fatty acids (arachidic and nonadecylic acid) and several metabolites related to niacin metabolism (trigonelline, N1-methyl-2-pyridone-5-carboxamide, N1-methyl-4-pyridone-3-carboxamide). Notably, only 3 metabolites overlapped with the previously developed 20-metabolite MDS, including serotonin, thiamine and LPE 18:0/0:0. Further, while γ-aminobutyric acid (GABA) was included in the original MDS but did not reach nominal significance in our sample of Black individuals, the GABA derivative, 4-acetamidobutanoic acid, was identified.

**Figure 2.**
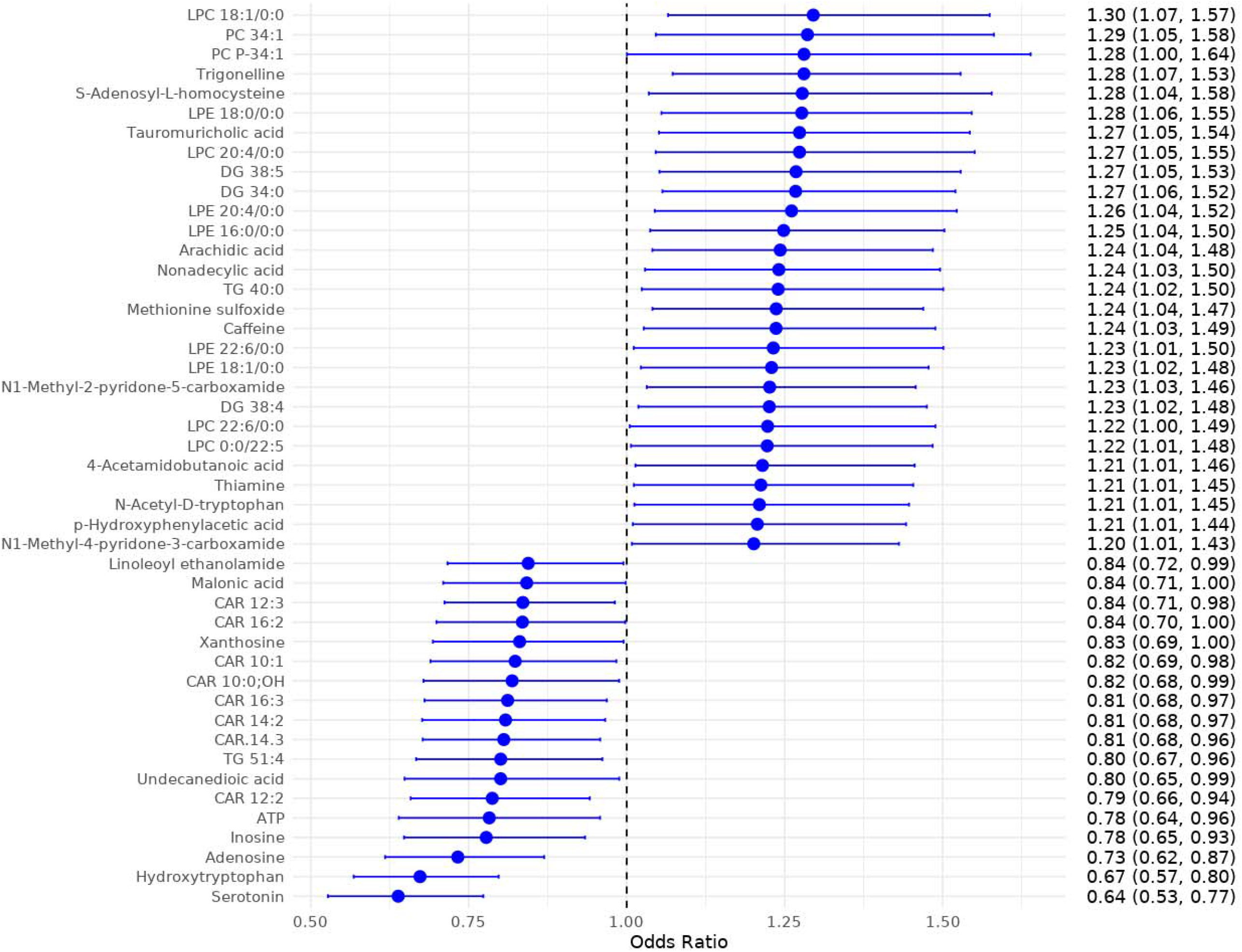
Metabolites (n=46) associated with depression status among Black MESA participants (n=1,140). Estimates represent the odds ratios for depression per one-unit increase in each metabolite z-score, adjusted for age, sex, study site, education, income, employment status, BMI, smoking status, physical activity, diet quality, systolic blood pressure, HDL cholesterol, total cholesterol, diabetes status, antihypertensive medications, and lipid-lowering medications.

### Stage 3: Development of the MDS-ME in MESA Black participants

LASSO selection was conducted based on 63 metabolites, including 46 nominally associated in Stage 2 in MESA Black participants and 20 previously identified in White participants (with 3 overlapping metabolites between the two sets). Of these, 33 metabolites had non-zero coefficients in LASSO selection, including 23 metabolites associated with depression in Black participants and 10 present in the original MDS (**Table 3**). The MDS-ME based on these 33 metabolites was correlated with a Pearson correlation of 0.33 (p<0.001) with the original MDS in this discovery sample of Black participants.

**Table 3.**
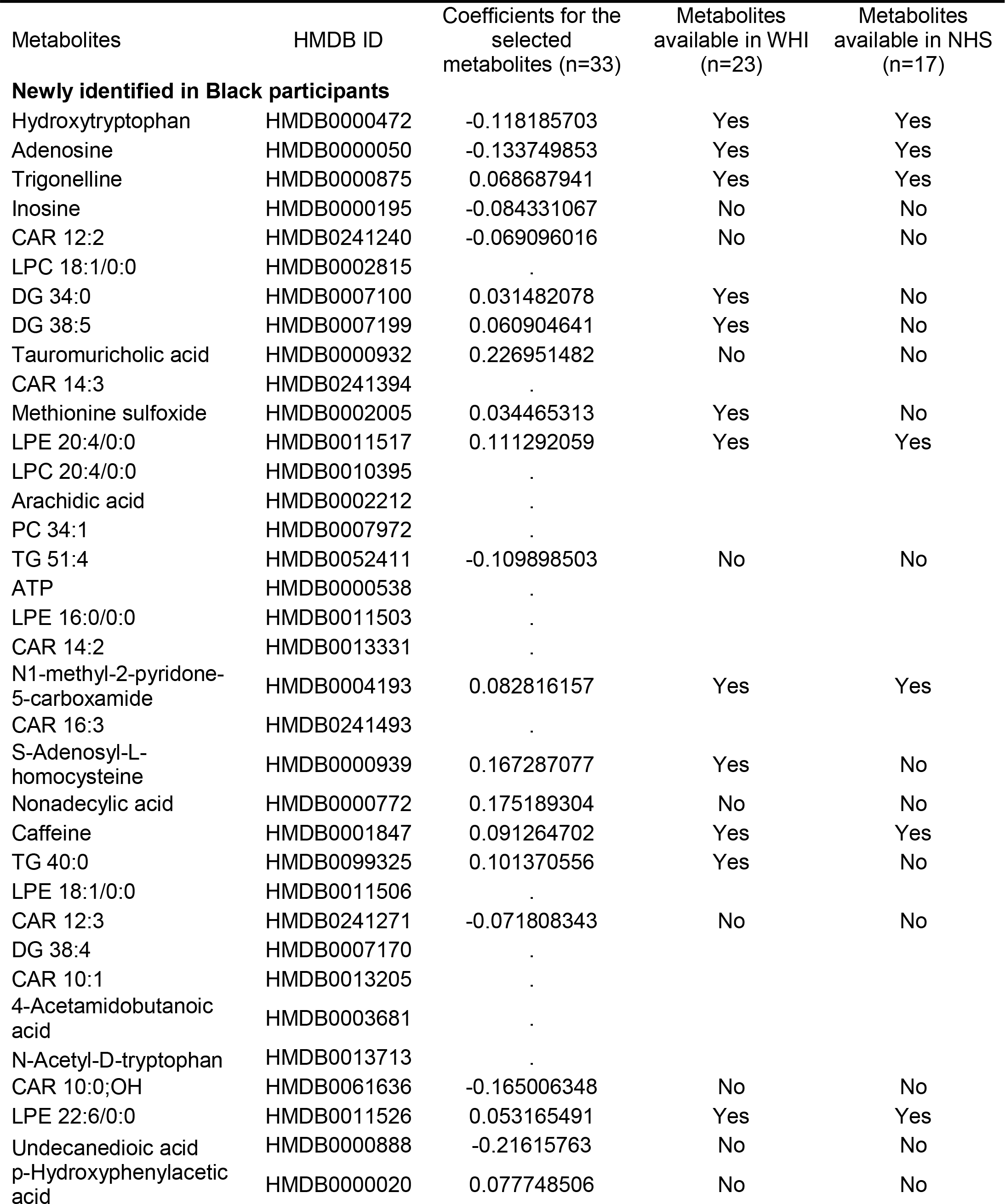
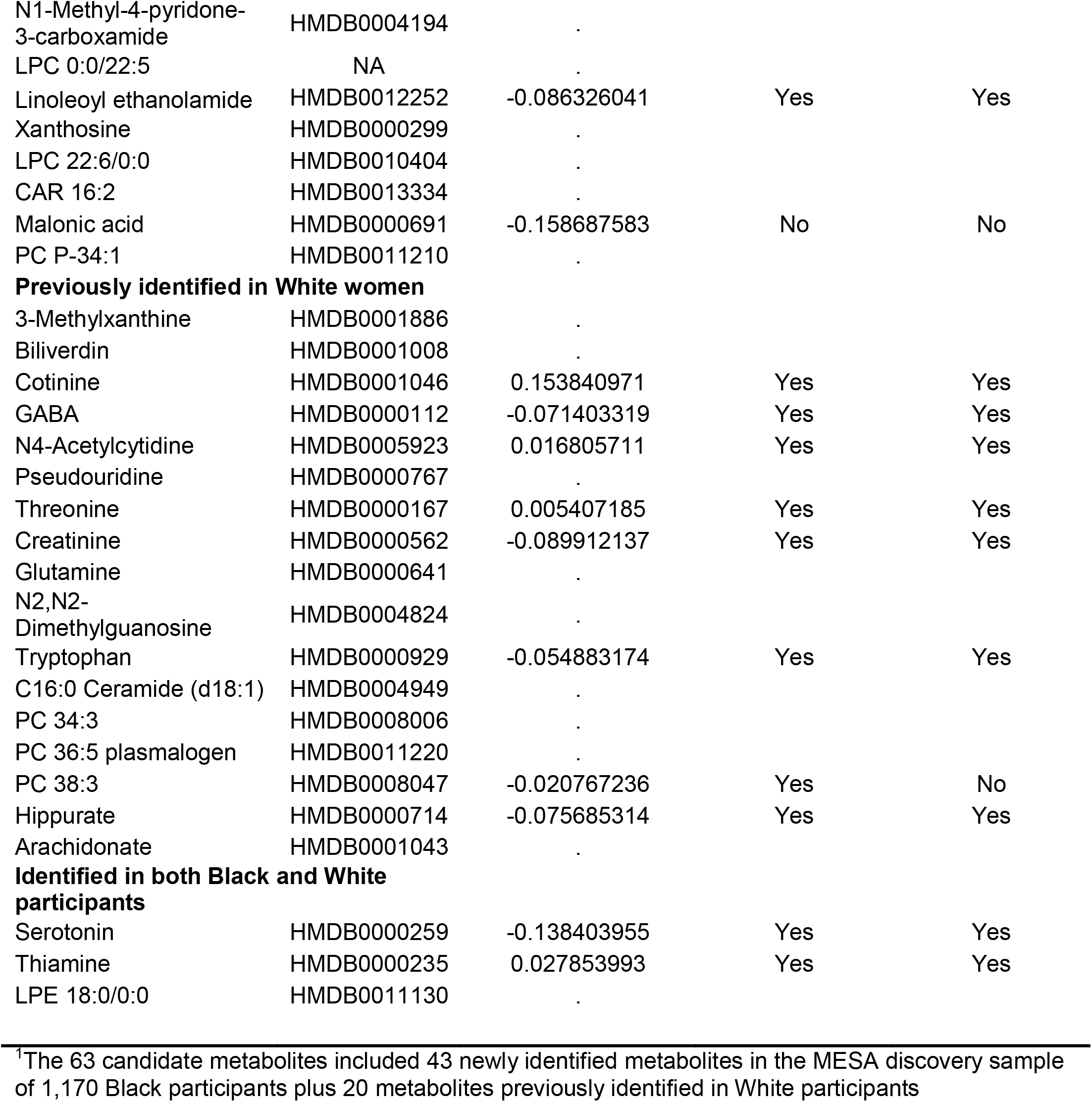
Metabolites selected by LASSO from 63 candidates^1^.

### Stage 4: Evaluation of the MDS-ME with depression in three independent samples

In the independent withheld replication sample of 485 MESA Black participants, the MDS-ME also had a Pearson correlation of 0.33 with the original MDS. After full adjustment in Model 2, the association between MDS-ME and depression was statistically significant (OR per 1-SD increment in the MDS-ME: 1.45, 95% CI: 1.04, 2.03), with no evidence for sex differences (p-interaction>0.78; **Table 4**). The MDS-ME also performed well among MESA White participants (n=2,477; fully-adjusted OR: 1.81; 95% CI: 1.60, 2.04), and the associations were similar in White men and women (p-interaction>0.49).

**Table 4.** Associations between the multi-ethnic MDS (per 1-SD increment) and depression status in White and Black participants from MESA.

| White and Black participants from MESA |  |  |  |  |  |
| --- | --- | --- | --- | --- | --- |
|  |  | Model 1<sup>1</sup> |  | Model 2<sup>2</sup> |  |
|  | n | OR (95% CI) | p-value | OR (95% CI) | p-value |
| MESA |  |  |  |  |  |
| Black participants (30% replication set) |  |  |  |  |  |
| Overall | 485 | 1.47 (1.09, 1.99) | 0.01 | 1.45 (1.04, 2.03) | 0.03 |
| Women | 264 | 1.43 (0.98, 2.13) | 0.07 | 1.46 (0.95, 2.27) | 0.09 |
| Men | 221 | 1.45 (0.89, 2.50) | 0.15 | 1.46 (0.79, 2.81) | 0.24 |
| p-int by sex |  | 0.94 |  | 0.78 |  |
| White participants |  |  |  |  |  |
| Overall | 2477 | 1.83 (1.63, 2.05) | 3.69E-25 | 1.81 (1.60, 2.04) | 2.67E-22 |
| Women | 1298 | 1.76 (1.52, 2.03) | 1.50E-14 | 1.75 (1.51, 2.04) | 2.02E-13 |
| Men | 1179 | 1.98 (1.64, 2.41) | 3.78E-12 | 1.94 (1.59, 2.39) | 2.39E-10 |
| p-int by sex |  | 0.49 |  | 0.58 |  |

<sup>1</sup>Adjusted for age, sex, study site, education, income, and employment status
<sup>2</sup>Model 1 + adjusted for BMI, smoking status, physical activity, diet quality, systolic blood pressure, HDL cholesterol, total cholesterol, diabetes status, antihypertensive medications, and lipid-lowering medications

In <u>WHI</u>, 23 out of the 33 metabolites selected by the LASSO in the MDS-ME developed in MESA were available; In <u>NHS</u>, 17 of 33 metabolites were available (**Table 3**). Among 485 MESA Black participants, the 33-metabolite MDS-ME was strongly correlated with the reduced 23-metabolite MDS-ME available in WHI (*r*=0.74) and with the reduced 17-metabolite MDS-ME available in NHS (*r*=0.64); the corresponding correlations were *r*=0.74 and *r*=0.65 among White MESA participants. These correlation levels suggested that using these reduced MDS-ME as proxies for the replication analyses is reasonable. In <u>WHI</u>, the reduced MDS-ME was associated with depression with a similar magnitude in White (n=1,107; OR: 1.21; 95% CI: 1.04, 1.40) and Black (n=149; OR: 1.34; 95% CI: 0.85, 2.21) women (**Table 5**). In <u>NHS</u>, the reduced MDS-ME was also associated with depression similarly in White (n=1,856; OR: 1.11; 95% CI: 0.95, 1.30) and Black (n=214; OR: 1.20; 95% CI: 0.69, 2.06) women. Notably, though the associations did not reach statistical significance among the two small samples of Black participants in WHI and NHS, we found no evidence for significant between-study heterogeneity based on the meta-analysis of fully-adjusted estimates in Black participants using the full MDS-ME in MESA, the 23-metabolite MDS-ME in WHI and the 17-metabolite MDS-ME in NHS (pooled OR: 1.36; 95% CI: 1.07, 1.74; I^2^=0%; p for heterogeneity=0.84; **Figure 3**).

**Figure 3.**
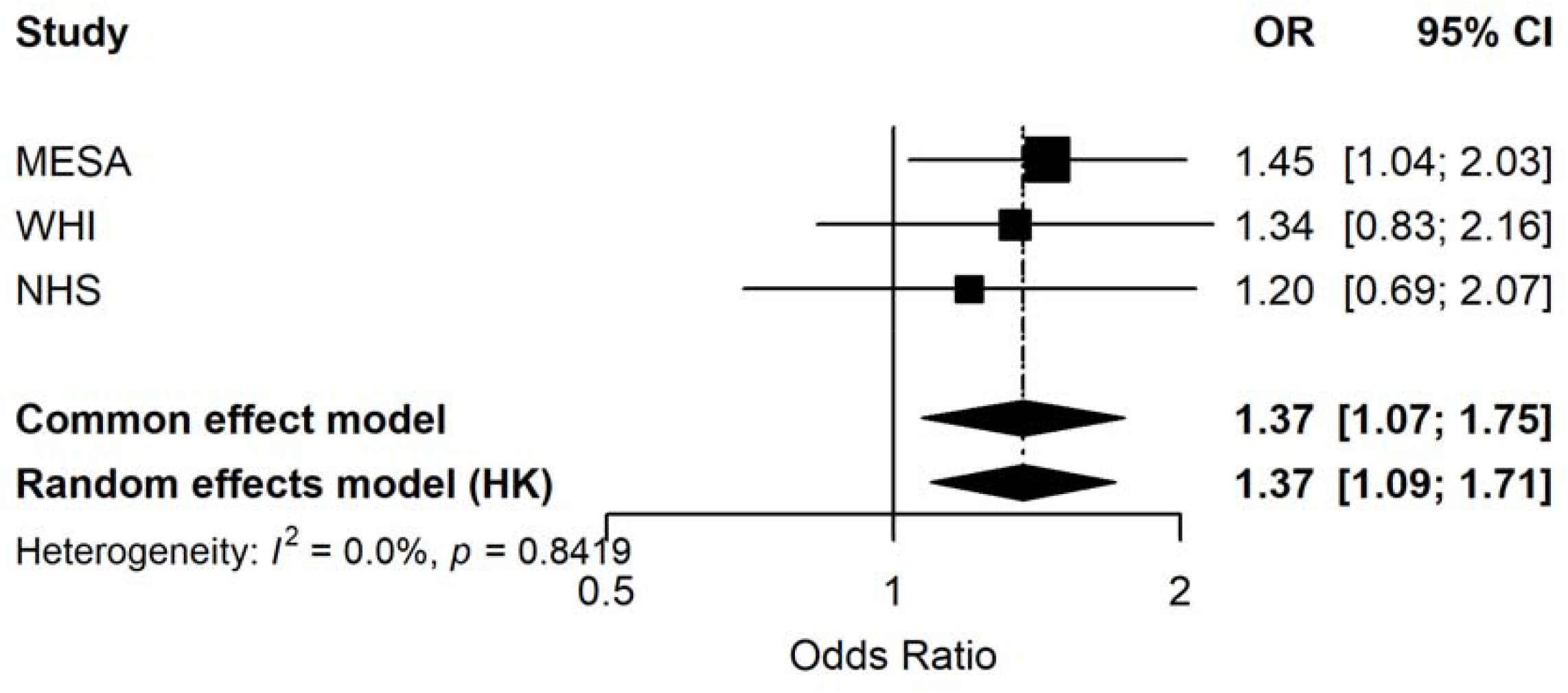
Meta-analysis of the association between the multi-ethnic MDS (MDS-ME) and depression among Black participants from MESA, WHI and NHS. The MDS-ME was calculated based on the full 33 metabolites in MESA, 23 metabolites in WHI and 17 metabolites in NHS.

**Table 5.**
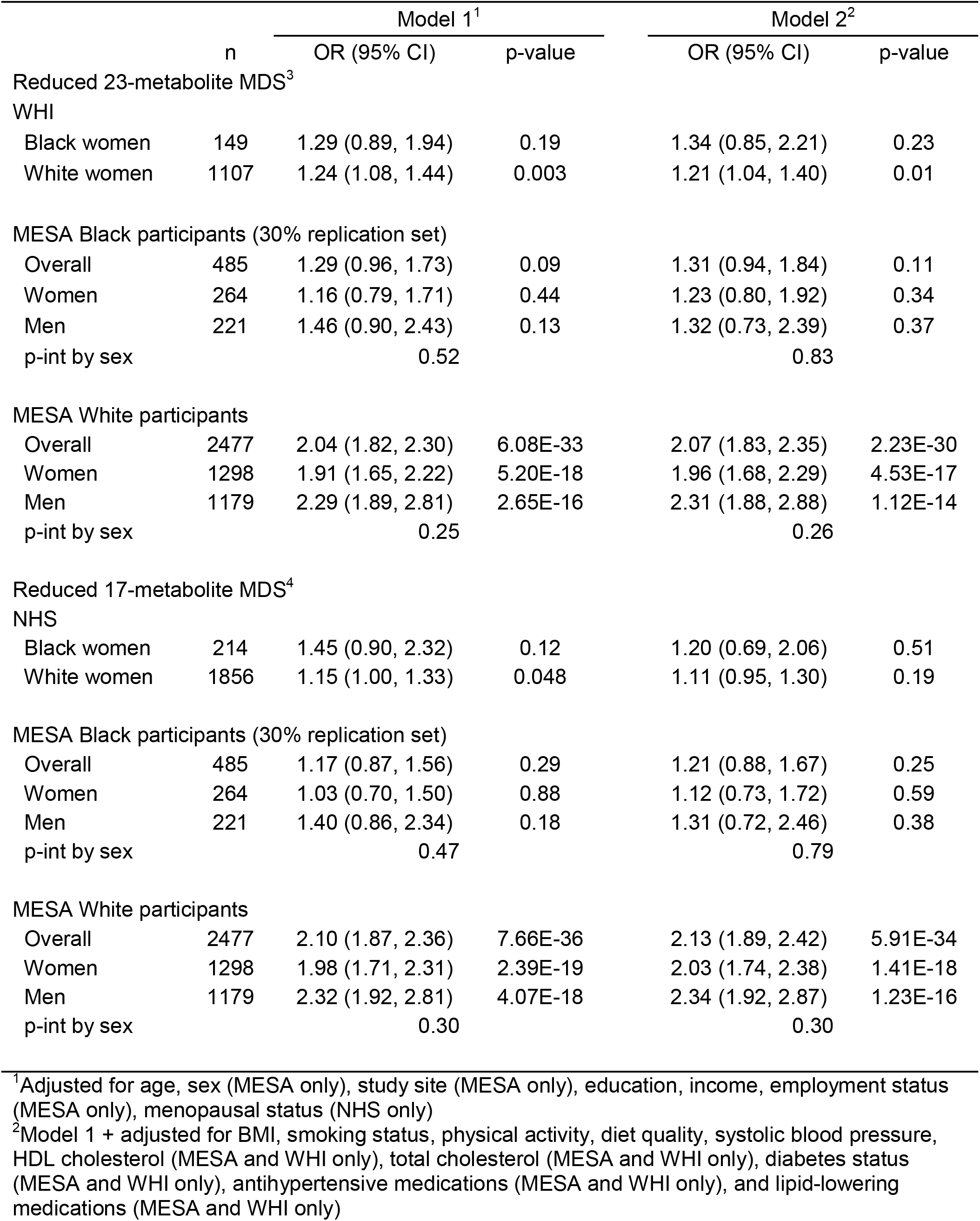
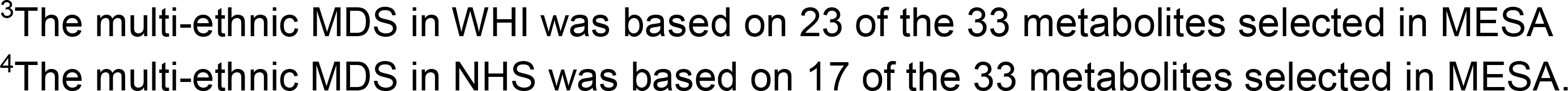
Associations between the reduced, multi-ethnic MDS (per 1-SD increment) and depression status in White and Black participants from WHI, NHS and MESA.

|  |  | Model 1<sup>1</sup> |  | Model 2<sup>2</sup> |  |
| --- | --- | --- | --- | --- | --- |
|  | n | OR (95% CI) | p-value | OR (95% CI) | p-value |
| Reduced 23-metabolite MDS<sup>3</sup> |  |  |  |  |  |
| WHI |  |  |  |  |  |
| Black women | 149 | 1.29 (0.89, 1.94) | 0.19 | 1.34 (0.85, 2.21) | 0.23 |
| White women | 1107 | 1.24 (1.08, 1.44) | 0.003 | 1.21 (1.04, 1.40) | 0.01 |
| MESA Black participants (30% replication set) |  |  |  |  |  |
| Overall | 485 | 1.29 (0.96, 1.73) | 0.09 | 1.31 (0.94, 1.84) | 0.11 |
| Women | 264 | 1.16 (0.79, 1.71) | 0.44 | 1.23 (0.80, 1.92) | 0.34 |
| Men | 221 | 1.46 (0.90, 2.43) | 0.13 | 1.32 (0.73, 2.39) | 0.37 |
| p-int by sex |  | 0.52 |  | 0.83 |  |
| MESA White participants |  |  |  |  |  |
| Overall | 2477 | 2.04 (1.82, 2.30) | 6.08E-33 | 2.07 (1.83, 2.35) | 2.23E-30 |
| Women | 1298 | 1.91 (1.65, 2.22) | 5.20E-18 | 1.96 (1.68, 2.29) | 4.53E-17 |
| Men | 1179 | 2.29 (1.89, 2.81) | 2.65E-16 | 2.31 (1.88, 2.88) | 1.12E-14 |
| p-int by sex |  | 0.25 |  | 0.26 |  |
| Reduced 17-metabolite MDS<sup>4</sup> |  |  |  |  |  |
| NHS |  |  |  |  |  |
| Black women | 214 | 1.45 (0.90, 2.32) | 0.12 | 1.20 (0.69, 2.06) | 0.51 |
| White women | 1856 | 1.15 (1.00, 1.33) | 0.048 | 1.11 (0.95, 1.30) | 0.19 |
| MESA Black participants (30% replication set) |  |  |  |  |  |
| Overall | 485 | 1.17 (0.87, 1.56) | 0.29 | 1.21 (0.88, 1.67) | 0.25 |
| Women | 264 | 1.03 (0.70, 1.50) | 0.88 | 1.12 (0.73, 1.72) | 0.59 |
| Men | 221 | 1.40 (0.86, 2.34) | 0.18 | 1.31 (0.72, 2.46) | 0.38 |
| p-int by sex |  | 0.47 |  | 0.79 |  |
| MESA White participants |  |  |  |  |  |
| Overall | 2477 | 2.10 (1.87, 2.36) | 7.66E-36 | 2.13 (1.89, 2.42) | 5.91E-34 |
| Women | 1298 | 1.98 (1.71, 2.31) | 2.39E-19 | 2.03 (1.74, 2.38) | 1.41E-18 |
| Men | 1179 | 2.32 (1.92, 2.81) | 4.07E-18 | 2.34 (1.92, 2.87) | 1.23E-16 |
| p-int by sex |  | 0.30 |  | 0.30 |  |

<sup>1</sup>Adjusted for age, sex (MESA only), study site (MESA only), education, income, employment status (MESA only), menopausal status (NHS only)
<sup>2</sup>Model 1 + adjusted for BMI, smoking status, physical activity, diet quality, systolic blood pressure, HDL cholesterol (MESA and WHI only), total cholesterol (MESA and WHI only), diabetes status (MESA and WHI only), antihypertensive medications (MESA and WHI only), and lipid-lowering medications (MESA and WHI only)
<sup>3</sup>The multi-ethnic MDS in WHI was based on 23 of the 33 metabolites selected in MESA
<sup>4</sup>The multi-ethnic MDS in NHS was based on 17 of the 33 metabolites selected in MESA.

In sensitivity analyses, as expected, the reduced MDS-ME showed weaker associations with depression than the full MDS-ME among MESA Black participants; however, the point estimates were remarkably similar to those observed in WHI and NHS Black women (**Table 5**). Moreover, both reduced MDS-ME demonstrated consistently strong positive associations with depression among MESA White participants.

## Discussion

In this study, we found that the MDS, originally developed in several independent samples of White women, was strongly associated with phenotypic measures of depression in both White men and women in MESA. In contrast, associations were weaker in Black participants, particularly Black women. We conducted an agnostic analysis to identify additional plasma metabolites associated with psychological distress in Black participants and revised the MDS by incorporating newly identified metabolites. The MDS-ME demonstrated stronger associations than the MDS with depression in Black men and women in MESA, as well as comparable associations in external samples of Black women from WHI and NHS. Nonetheless, the magnitude of the association between the MDS-ME and depression remained weaker in Black participants compared with that in White participants in the MESA sample.

Of note, the MDS-ME, developed among Black men and women in combination, was as strongly linked to depression in White men and women as the original MDS developed specifically among White women. These findings have several important implications. When examining metabolomic processes and their relationship to distress (or other psychosocial factors), it is important to consider potential racial differences and recognize that the relevant phenotypes may be heterogeneous across populations. Also important to recognize is that more heterogeneous samples may provide greater insight into how metabolomic processes are associated with higher order factors. Similar themes have emerged in psychiatric genetics, where multi-ancestry studies of major depression have identified novel loci and shown that newer polygenic score methods that integrate ancestry-diverse GWAS data improve cross-population prediction (2025; Hoggart et al., 2024; Meng et al., 2024; Ruan et al., 2022). Likewise, studies of plasma metabolomics have documented substantial Black-White differences, particularly in lipids and amino acids (Butler et al., 2023; Hu et al., 2022; McGee et al., 2024). In women, a metabolomic pattern capturing such Black-White differences was associated with incident coronary heart disease risk independent of self-reported race and traditional risk factors (Hu et al., 2022), underscoring the value of more heterogeneous samples for clarifying how distress-related metabolomic signatures may contribute to health disparities.

Among Black participants, the three metabolites that were most strongly associated with depression status and remained significant after correction for multiple comparisons were serotonin, hydroxytryptophan, and adenosine. Hydroxytryptophan is a natural intermediate product on the pathway that converts tryptophan to serotonin, a key neurotransmitter involved in stress responses, mood regulation, and sleep (Maffei, 2020). Because hydroxytryptophan directly influences serotonin synthesis, altered serotonin signaling due to changes in hydroxytryptophan levels or receptor functions have been implicated in depressive symptoms (Correia and Vale, 2022; Richard et al., 2009). Adenosine is known to have a predominantly inhibitory effect on the central nervous system. Reduced adenosine levels have been associated with elevated chronic inflammation, dysregulated dopamine and glutamate activity, and exaggerated stress responses, all of which may increase vulnerability to depression (Haskó and Cronstein, 2013; Pasquini et al., 2022; Yue et al., 2025). This is consistent with our observation of an inverse association between adenosine and depression. Interestingly, we also observed a positive association of caffeine with depression. Caffeine primarily acts as an adenosine receptor antagonist in increasing alertness, enhancing concentration, and increasing wakefulness (the opposite of symptoms found in depression) (Ribeiro and Sebastião, 2010). Given that caffeine metabolism may vary across individuals and may be influenced by broader contextual factors (Rybak et al., 2015), further investigation is required to fully elucidate the complex biological consequences of the balance between adenosine, caffeine, and adenosine receptor across populations.

Other metabolites associated with depression among Black participants included positive associations for several lipid species and inverse associations for multiple carnitines. Changes in lipid profiles, such as increased levels of PCs, LPCs, and LPEs, may be linked to depression by disrupting cell-membrane integrity, neurotransmitter signaling, inflammation, and mitochondrial energy metabolism, which can impair neuronal function and stress regulation (Ge et al., 2025; Pinto et al., 2022). Conversely, carnitines tend to show the opposite metabolic patterns by facilitating fatty-acid transport into mitochondria for increased mitochondrial efficiency, enhancing neuroplasticity, and exerting anti-inflammatory effects, with lower levels indicating impaired energy metabolism and diminished adaptive neural responses commonly seen in depression (Ait Tayeb et al., 2024; Bigio et al., 2024; Milaneschi et al., 2022). Thus, while altered lipids often signal metabolic stress and inflammation, lower carnitine levels suggest mitochondrial underperformance as a complementary biological pathway in depression. In addition, thiamine (vitamin B1) and several metabolites related to niacin (vitamin B3) metabolism were positively associated with depression status. It is possible that increased niacin, which is synthesized from tryptophan, may reduce tryptophan bioavailability for serotonin production, thereby altering serotonin signaling and increasing depression risk (Davidson et al., 2022; Tang et al., 2025). However, some evidence suggests that vitamin B deficiency may also contribute to depressive symptoms (Han et al., 2025; Mikkelsen et al., 2017). Given the conflicting evidence, the role of niacin-related metabolites in depression warrants further study.

Of note, many of the metabolites discussed above based on findings in Black participants had not shown the most robust signals with depression in our previous study of White women. In fact, only three metabolites (serotonin, thiamine and LPE 18:0/0:0) previously included in the MDS developed in White women were significantly associated with depression in MESA Black participants; several others, including GABA, did not reach nominal significance. Such differences in the distress-related metabolic profiles between White and Black participants may reflect underlying variation in genetic architecture, metabolic regulation, or inflammatory responses across populations. Socio-environmental factors, such as unemployment, discrimination, diet, and neighborhood context, may also influence metabolic profiles in ways that intersect with depression risk (Cardel et al., 2021; Li et al., 2019; Vrany et al., 2021). Additionally, unequal access to treatment and differences in comorbid conditions could influence both metabolic pathways and depression. Collectively, these factors likely interact, leading to different metabolomic signatures of depression that emerge as important in diverse groups.

Despite our efforts to refine the MDS for Black participants, the strength of its association with depression among Black participants with MDS was still weaker than associations among White participants using either the original MDS or the MDS-ME. Methodologically, a more limited sample size for Black participants may have reduced power for metabolite discovery and replication, potentially attenuating observed associations. Biologically, our findings may suggest that depression, or more generally psychological distress, is a more heterogenous phenotype in Black participants, influenced by a broad range of social and environmental risk factors (Assari, 2017; Assari and Moazen-Zadeh, 2016; Vyas et al., 2020; Wang and Narcisse, 2025). If future studies confirm reliable differences, this could suggest caution in generalizing biomarker-based screening tools developed in homogeneous samples. At the population level, these findings highlight the importance of considering how structural and social determinants of health can affect metabolic processes associated with psychological distress in diverse groups.

This study has several key strengths. First, it focused on two populations at high risk for cardiometabolic disease, Black individuals and men, to identify metabolomic signatures associated with psychological distress. Second, a rigorous approach was applied to revise the MDS and replicate the associations both internally and externally in independent samples. Third, a common, extensively validated metabolomic profiling method was used across datasets, facilitating direct comparison. Finally, key covariates were available across different samples, allowing for comprehensive and comparable adjustment in all analyses.

Several limitations should also be noted. Our study was cross-sectional by design, and the identified metabolomic profiles may reflect both metabolic precursors and consequences of depression. Longitudinal studies and causal approaches (e.g., Mendelian randomization) are needed to disentangle these potentially bidirectional relationships. While our study represents the first and largest investigation on metabolomic profiles of psychological distress specifically for Black individuals, the sample size for Black participants was still modest, particularly compared with that for White participants. This may have limited our ability to identify certain metabolites with smaller effect sizes. Despite consistency in metabolomic profiling and covariate assessment, there were variations in the definition of depression and population characteristics (e.g., age and sex distributions) across study samples. Also, our analysis was limited to metabolites consistently annotated across independent cohorts. Because data processing pipelines and annotation approaches have advanced significantly over time, older cohorts contain more unannotated features that, if identified, could have strengthened validation or further improved the MDS-ME. Furthermore, the influence of long-term and variable blood sample storage on plasma metabolites is not well understood (Townsend et al., 2016; Yin et al., 2015). These could compromise replication efforts and reduce the generalizability of the findings to contemporary populations. Future studies with robust replication are also needed for other racial and ethnic groups (e.g., Asians, Hispanics) to develop a comprehensive understanding of the metabolic correlates of psychological distress in diverse populations.

In summary, the MDS originally developed in White women showed similar associations with depression in White men but weaker associations in Black individuals. Incorporating additional metabolites associated with depression in Black individuals yielded MDS-ME that showed more consistent positive associations with depression status across three independent replication samples of Black individuals, highlighting a broader array of metabolites that may underlie depression than was previously identified. However, the association between the MDS-ME and depression in Black individuals remained weaker than that between the original MDS and depression in White individuals, suggesting additional metabolites may emerge with larger and more diverse samples. Future studies should extend investigations of distress-related metabolic profiles to larger and more diverse populations and assess how such profiles contribute to cardiometabolic and other health outcomes and if they provide insight into health disparities across diverse groups.

## Data Availability

All data produced in the present work are contained in the manuscript

## Acknowledgements

We thank Dr. Robert Gerszten for his contribution to the generation of the metabolomics datasets used in this study.

This study is supported by the National Institutes of Health Grant R01AG051600. The content is solely the responsibility of the authors and does not necessarily represent the official views of the National Institutes of Health. TH is supported by the Intramural Research Program of the National Institutes of Health, National Institute on Aging (ZIAAG000530). ACW is supported, in part, by USDA/ARS (Cooperative Agreement 58-3092-5-001). The contents of this publication do not necessarily reflect the views or policies of the USDA, nor does mention of trade names, commercial products, or organizations imply endorsement from the US government.

The Multi-Ethnic Study of Atherosclerosis is supported by contracts 75N92025D00022, 75N92020D00001,HHSN268201500003I, N01-HC-95159, 75N92025D00026, 75N92020D00005, N01-HC-95160,75N92020D00002, N01-HC-95161, 75N92025D00024, 75N92020D00003, N01-HC-95162,75N92025D00027, 75N92020D00006, N01-HC-95163, 75N92025D00025, 75N92020D00004, N01-HC-95164,75N92025D00028, 75N92020D00007, N01-HC-95165, N01-HC-95166, N01-HC-95167, N01-HC-95168, N01-HC-95169, UL1-TR-000040, UL1-TR-001079, UL1-TR-001420 from the National Center for Advancing Translational Sciences (NCATS). The authors thank the other investigators, the staff, and the participants of the MESA study for their valuable contributions. A full list of participating MESA investigators and institutions can be found at http://www.mesa-nhlbi.org. This manuscript has been reviewed by MESA for scientific content and consistency of data interpretation with previous MESA publications.

Molecular data for the Trans-Omics for Precision Medicine program were supported by the National Heart, Lung, and Blood Institute. Metabolomics data for MESA (phs001416) were generated at the Broad Institute and Beth Israel Metabolomics Platform (HHSN268201600038I) during TOPMed phases pilot, 5.5, and 8. Core support including centralized genomic read mapping and genotype calling, along with variant quality metrics and filtering were provided by the TOPMed Informatics Research Center (3R01HL-117626-02S1; contract HHSN268201800002I). Core support including phenotype harmonization, data management, sample-identity QC, and general program coordination were provided by the TOPMed Data Coordinating Center (R01HL-120393; U01HL-120393; contract HHSN268201800001I). We gratefully acknowledge the studies and participants who provided biological samples and data for TOPMed.

The WHI program is funded by the National Heart, Lung, and Blood Institute, National Institutes of Health, U.S. Department of Health and Human Services through contracts 75N92021D00001, 75N92021D00002, 75N92021D00003, 75N92021D00004, 75N92021D00005. Metabolomic analysis in the WHI was funded by the National Heart, Lung, and Blood Institute, National Institutes of Health, U.S. Department of Health and Human Services through contract HHSN268201300008C. The authors thank the WHI investigators and staff for their dedication, and the study participants for making the program possible. A listing of WHI investigators can be found at https://www.whi.org/doc/WHIInvestigator-Long-List.pdf. This manuscript was prepared in collaboration with investigators of the WHI, and has been reviewed and/or approved by the Women’s Health Initiative (WHI).

We gratefully acknowledge the Nurses’ Health Study (NHS) and Nurses’ Health Study II (NHSII) participants and staff for their invaluable contributions. This project was supported by NIH grants UM1 CA186107 (NHS), U01 CA176726 (NHSII), R01 CA49449, and R01 HL034594. The content is solely the responsibility of the authors and does not necessarily represent the official views of the National Institutes of Health.

---

**Supplemental Table 1.**
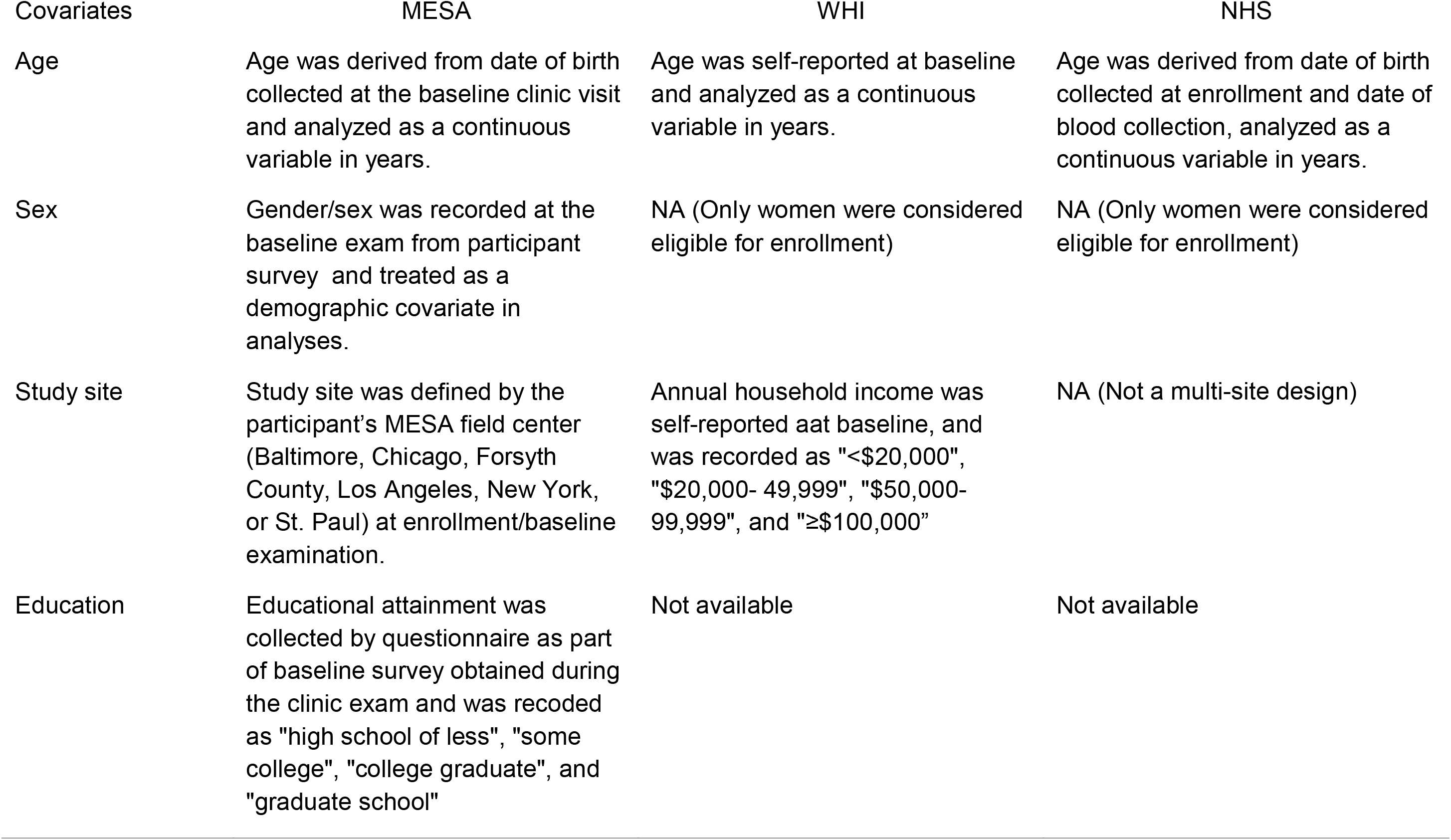
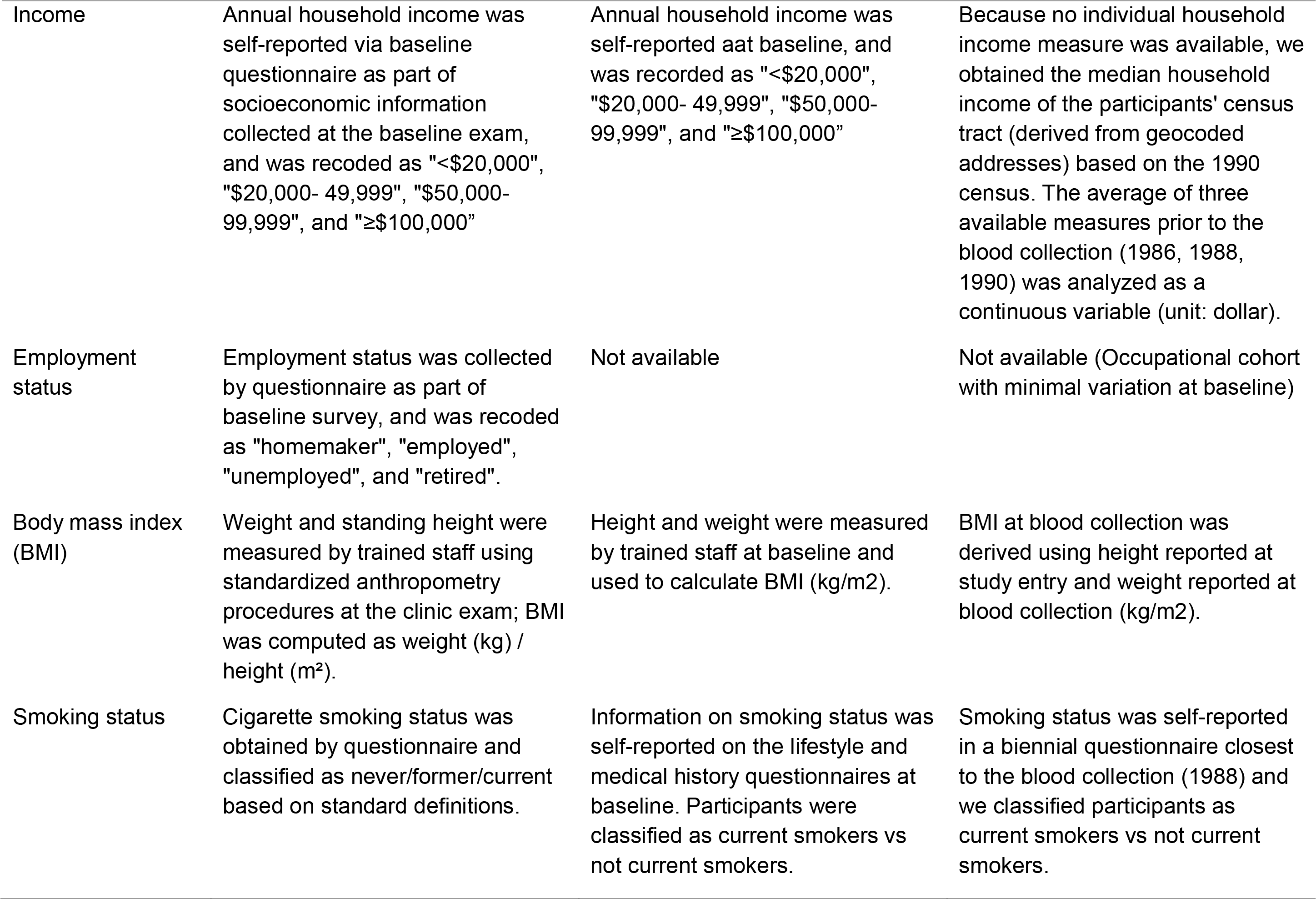
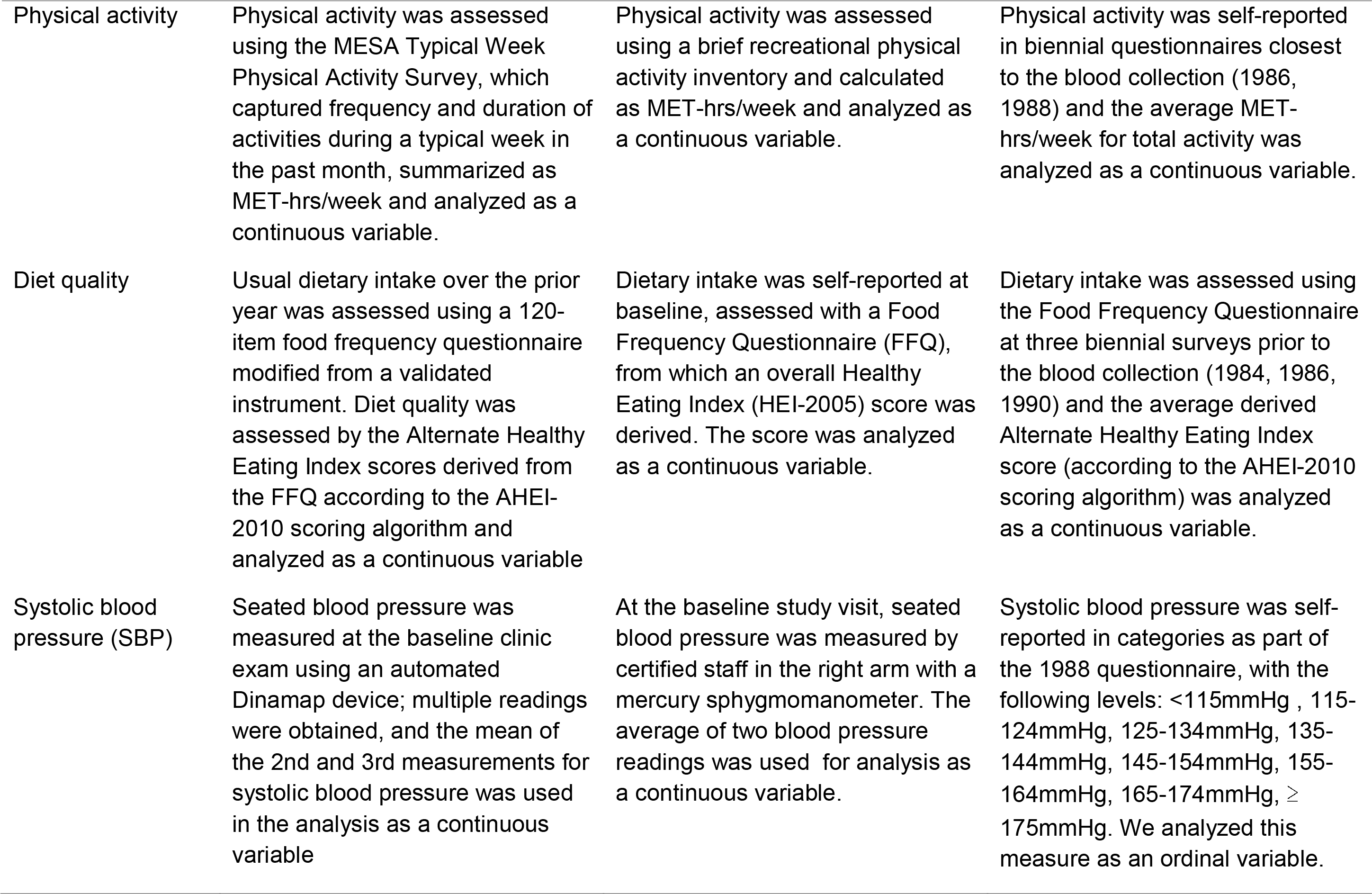
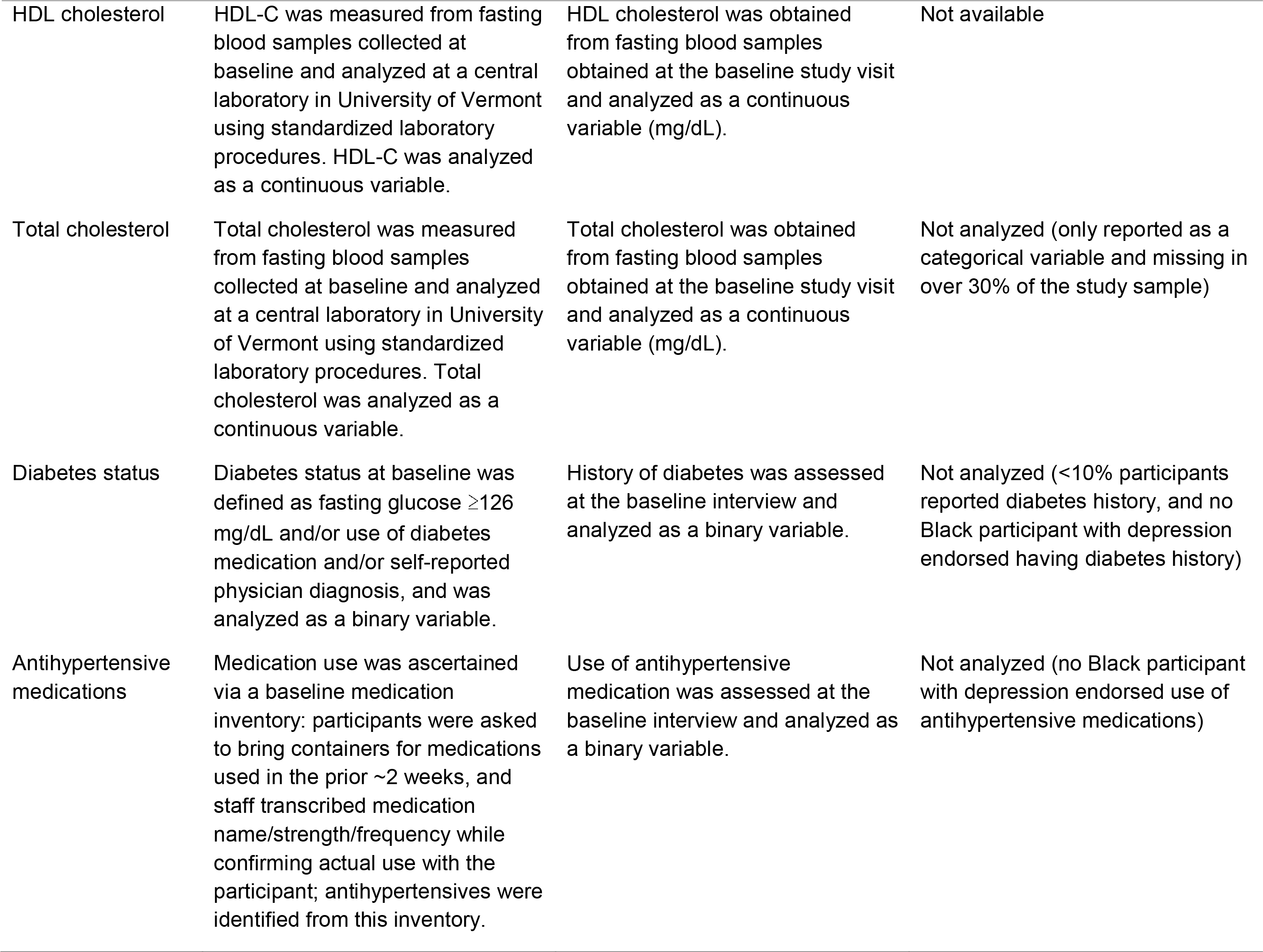
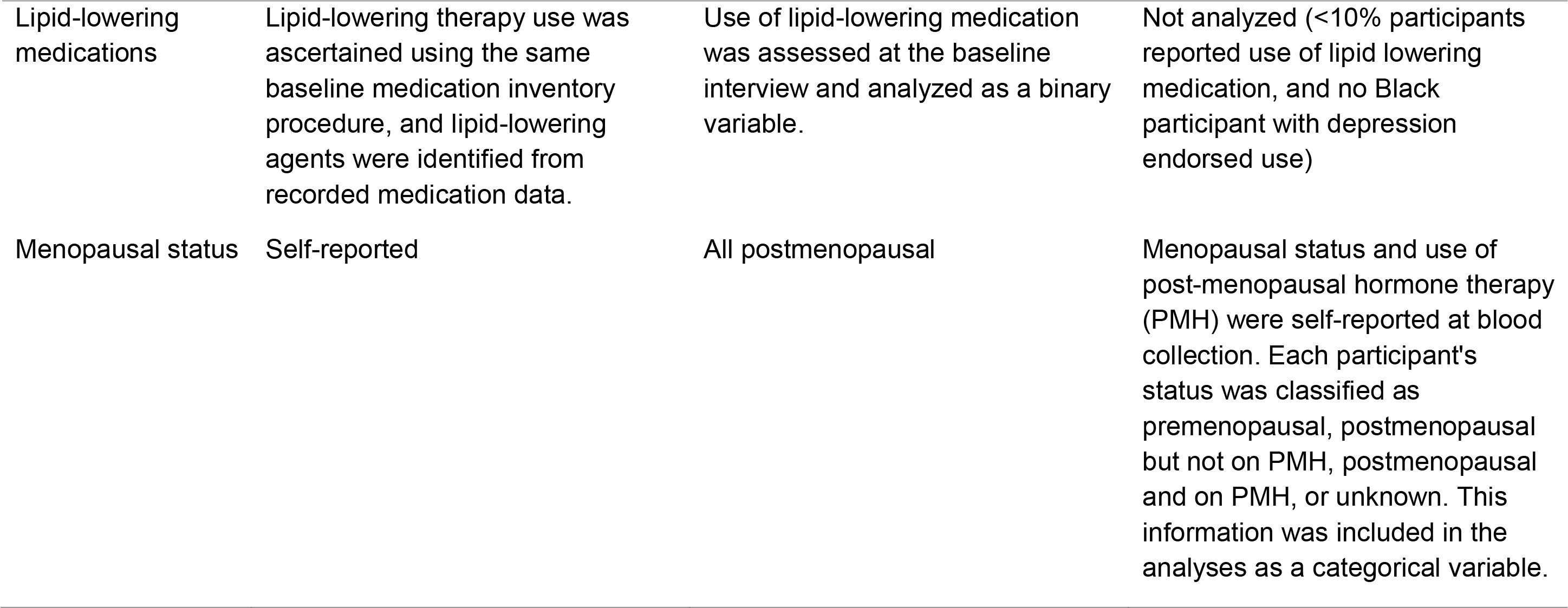
Covariate assessment in each participating cohort.

| Covariates | MESA | WHI | NHS |
| --- | --- | --- | --- |
| Age | Age was derived from date of birth collected at the baseline clinic visit and analyzed as a continuous variable in years. | Age was self-reported at baseline and analyzed as a continuous variable in years. | Age was derived from date of birth collected at enrollment and date of blood collection, analyzed as a continuous variable in years. |
| Sex | Gender/sex was recorded at the baseline exam from participant survey and treated as a demographic covariate in analyses. | NA (Only women were considered eligible for enrollment) | NA (Only women were considered eligible for enrollment) |
| Study site | Study site was defined by the participant's MESA field center (Baltimore, Chicago, Forsyth County, Los Angeles, New York, or St. Paul) at enrollment/baseline examination. | Annual household income was self-reported at baseline, and was recorded as "<\$20,000", "\$20,000- 49,999", "\$50,000- 99,999", and "≥\$100,000" | NA (Not a multi-site design) |
| Education | Educational attainment was collected by questionnaire as part of baseline survey obtained during the clinic exam and was recoded as "high school or less", "some college", "college graduate", and "graduate school" | Not available | Not available |
| Income | Annual household income was self-reported via baseline questionnaire as part of socioeconomic information collected at the baseline exam, and was recoded as "<\$20,000", "\$20,000- 49,999", "\$50,000- 99,999", and "≥\$100,000" | Annual household income was self-reported at baseline, and was recorded as "<\$20,000", "\$20,000- 49,999", "\$50,000- 99,999", and "≥\$100,000" | Because no individual household income measure was available, we obtained the median household income of the participants' census tract (derived from geocoded addresses) based on the 1990 census. The average of three available measures prior to the blood collection (1986, 1988, 1990) was analyzed as a continuous variable (unit: dollar). |
| Employment status | Employment status was collected by questionnaire as part of baseline survey, and was recoded as "homemaker", "employed", "unemployed", and "retired". | Not available | Not available (Occupational cohort with minimal variation at baseline) |
| Body mass index (BMI) | Weight and standing height were measured by trained staff using standardized anthropometry procedures at the clinic exam; BMI was computed as weight (kg) / height (m<sup>2</sup>). | Height and weight were measured by trained staff at baseline and used to calculate BMI (kg/m<sup>2</sup>). | BMI at blood collection was derived using height reported at study entry and weight reported at blood collection (kg/m<sup>2</sup>). |
| Smoking status | Cigarette smoking status was obtained by questionnaire and classified as never/former/current based on standard definitions. | Information on smoking status was self-reported on the lifestyle and medical history questionnaires at baseline. Participants were classified as current smokers vs not current smokers. | Smoking status was self-reported in a biennial questionnaire closest to the blood collection (1988) and we classified participants as current smokers vs not current smokers. |
| Physical activity | Physical activity was assessed using the MESA Typical Week Physical Activity Survey, which captured frequency and duration of activities during a typical week in the past month, summarized as MET-hrs/week and analyzed as a continuous variable. | Physical activity was assessed using a brief recreational physical activity inventory and calculated as MET-hrs/week and analyzed as a continuous variable. | Physical activity was self-reported in biennial questionnaires closest to the blood collection (1986, 1988) and the average MET-hrs/week for total activity was analyzed as a continuous variable. |
| Diet quality | Usual dietary intake over the prior year was assessed using a 120-item food frequency questionnaire modified from a validated instrument. Diet quality was assessed by the Alternate Healthy Eating Index scores derived from the FFQ according to the AHEI-2010 scoring algorithm and analyzed as a continuous variable | Dietary intake was self-reported at baseline, assessed with a Food Frequency Questionnaire (FFQ), from which an overall Healthy Eating Index (HEI-2005) score was derived. The score was analyzed as a continuous variable. | Dietary intake was assessed using the Food Frequency Questionnaire at three biennial surveys prior to the blood collection (1984, 1986, 1990) and the average derived Alternate Healthy Eating Index score (according to the AHEI-2010 scoring algorithm) was analyzed as a continuous variable. |
| Systolic blood pressure (SBP) | Seated blood pressure was measured at the baseline clinic exam using an automated Dinamap device; multiple readings were obtained, and the mean of the 2nd and 3rd measurements for systolic blood pressure was used in the analysis as a continuous variable | At the baseline study visit, seated blood pressure was measured by certified staff in the right arm with a mercury sphygmomanometer. The average of two blood pressure readings was used for analysis as a continuous variable. | Systolic blood pressure was self-reported in categories as part of the 1988 questionnaire, with the following levels: <115mmHg , 115-124mmHg, 125-134mmHg, 135-144mmHg, 145-154mmHg, 155-164mmHg, 165-174mmHg, ≥ 175mmHg. We analyzed this measure as an ordinal variable. |
| HDL cholesterol | HDL-C was measured from fasting blood samples collected at baseline and analyzed at a central laboratory in University of Vermont using standardized laboratory procedures. HDL-C was analyzed as a continuous variable. | HDL cholesterol was obtained from fasting blood samples obtained at the baseline study visit and analyzed as a continuous variable (mg/dL). | Not available |
| Total cholesterol | Total cholesterol was measured from fasting blood samples collected at baseline and analyzed at a central laboratory in University of Vermont using standardized laboratory procedures. Total cholesterol was analyzed as a continuous variable. | Total cholesterol was obtained from fasting blood samples obtained at the baseline study visit and analyzed as a continuous variable (mg/dL). | Not analyzed (only reported as a categorical variable and missing in over 30% of the study sample) |
| Diabetes status | Diabetes status at baseline was defined as fasting glucose $\geq 126$ mg/dL and/or use of diabetes medication and/or self-reported physician diagnosis, and was analyzed as a binary variable. | History of diabetes was assessed at the baseline interview and analyzed as a binary variable. | Not analyzed (<10% participants reported diabetes history, and no Black participant with depression endorsed having diabetes history) |
| Antihypertensive medications | Medication use was ascertained via a baseline medication inventory: participants were asked to bring containers for medications used in the prior ~2 weeks, and staff transcribed medication name/strength/frequency while confirming actual use with the participant; antihypertensives were identified from this inventory. | Use of antihypertensive medication was assessed at the baseline interview and analyzed as a binary variable. | Not analyzed (no Black participant with depression endorsed use of antihypertensive medications) |
| Lipid-lowering medications | Lipid-lowering therapy use was ascertained using the same baseline medication inventory procedure, and lipid-lowering agents were identified from recorded medication data. | Use of lipid-lowering medication was assessed at the baseline interview and analyzed as a binary variable. | Not analyzed (<10% participants reported use of lipid lowering medication, and no Black participant with depression endorsed use) |
| Menopausal status | Self-reported | All postmenopausal | Menopausal status and use of post-menopausal hormone therapy (PMH) were self-reported at blood collection. Each participant's status was classified as premenopausal, postmenopausal but not on PMH, postmenopausal and on PMH, or unknown. This information was included in the analyses as a categorical variable. |

**Supplemental Table 2.** List of 20 metabolites previously associated with depression in White women.

| Metabolites | HMDB ID | Weights |
| --- | --- | --- |
| 3-methylxanthine | HMDB01886 | 0.329765903 |
| biliverdin | HMDB01008 | -0.423583685 |
| cotinine | HMDB01046 | 0.366219583 |
| GABA | HMDB00112 | -0.152260834 |
| N4-acetylcytidine | HMDB05923 | -0.11936796 |
| pseudouridine | HMDB00767 | 0.049966016 |
| serotonin | HMDB00259 | -0.586268696 |
| threonine | HMDB00167 | -0.436917435 |
| creatinine | HMDB00562 | -0.358052338 |
| glutamine | HMDB00641 | 0.372924654 |
| N2,N2-dimethylguanosine | HMDB04824 | 0.574745872 |
| thiamine | HMDB00235 | 0.046196797 |
| tryptophan | HMDB00929 | -0.246402337 |
| C16:0 Ceramide (d18:1) | HMDB04949 | 0.129344695 |
| C18:0 LPE | HMDB11130 | 0.1123545 |
| C34:3 PC | HMDB08006 | -0.400454449 |
| C36:5 PC plasmalogen-B | HMDB11220 | -0.1138951 |
| C38:3 PC | HMDB08047 | 0.307754297 |
| hippurate | HMDB00714 | -0.425864438 |
| arachidonate | HMDB01043 | -0.329996729 |

**Supplemental Table 3.** Sample characteristics in White and Black women from WHI and NHS.

|  | WHI |  | NHS |  |
| --- | --- | --- | --- | --- |
|  | White | Black | White | Black |
| N | 1,107 | 149 | 1856 | 214 |
| CESD | 2.3 (2.5) | 2.9 (2.6) | - | - |
| CESD≥16, % | 15.3 | 20.8 | - | - |
| MHI-5 | - | - | 78.0 (13.5) | 77.8 (14.0) |
| MHI-5 ≤52, % | - | - | 5.6 | 7.3 |
| Antidepressant use, % | 7.1 | 4.7 | 6.4 | 2.8 |
| History of depressed mood, % | 10.8 | 15.4 | - | - |
| Depression diagnosis, % | - | - | 5.5 | 3.3 |
| Depression status, % | 24.1 | 28.9 | 12.4 | 10.3 |
| **Sociodemographic factors** |  |  |  |  |
| Age, yrs | 67.2 (6.8) | 64.2 (6.8) | 55.6 (6.9) | 56.1 (6.4) |
| Household income, % |  |  |  |  |
| <\$20,000 | 28.9 | 55.0 | - | - |
| \$20,000-\$49,999 | 52.1 | 36.2 | - | - |
| \$50,000-\$99,999 | 15.8 | 7.4 | - | - |
| ≥\$100,000 | 3.2 | 1.3 | - | - |
| Census tract-based median household income, \$ | - | - | 46766 (17087) | 39580 (16563) |
| **Medical factors** |  |  |  |  |
| Menopausal status and postmenopausal hormone (PMH) use, % |  |  |  |  |
| Premenopausal | - | - | 25.6 | 15.9 |
| Postmenopausal but not on PMH | - | - | 30.7 | 50.9 |
| Postmenopausal, on PMH | - | - | 31.2 | 22.9 |
| Unknown status | - | - | 12.4 | 10.3 |
| Systolic blood pressure | 132.8 (18.3) | 138.3 (20.6) | - | - |
| Self-reported systolic blood pressure categories |  |  |  |  |
| < 115 mmHg | - | - | 24.8 | 18.6 |
| 115 - 124 mmHg | - | - | 26.8 | 22.6 |
| 125 - 134 mmHg | - | - | 30.6 | 30.7 |
| 135 - 144 mmHg | - | - | 11.3 | 18.1 |
| 145 - 154 mmHg | - | - | 5.1 | 8.0 |
| 155 - 164 mmHg | - | - | 1.0 | 1.5 |
| 165 - 174 mmHg | - | - | 0.2 | 0.5 |
| ≥ 175 mmHg | - | - | 0.2 | 0 |
| HDL cholesterol, mg/dL | 52.3 (13.2) | 48.9 (12.4) | - | - |
| Total cholesterol, mg/dL | 235.5 (48.4) | 237.3 (40.0) | - | - |
| Diabetes status, %<sup>1</sup> | 22.4 | 13.5 | 3.0 | 6.5 |
| Antihypertensive medications, %<sup>1</sup> | 46.8 | 27.6 | 13.0 | 20.6 |
| Lipid-lowering medications, %<sup>1</sup> | 15.4 | 15.6 | 2.2 | 2.8 |
| **Lifestyle factors** |  |  |  |  |
| BMI | 29.0 (5.8) | 31.6 (6.2) | 25.3 (4.5) | 27.3 (5.3) |
| Current smokers, % | 13.8 | 16.0 | 12.8 | 13.6 |
| Physical activity, MET-hrs/week | 9.6 (11.1) | 8.7 (15.3) | 16.4 (20.1) | 13.0 (14.1) |
| Diet quality score | 66.8 (10.6) | 61.7 (11.1) | 51.8 (10.0) | 54.4 (9.6) |

<sup>1</sup>While information on diabetes, antihypertensive medications, and lipid-lowering medications was available in NHS, we were unable to include them as covariates in the analysis because among the small sample of Black participants, no participant with depression endorsed these variables

**Supplemental Table 4.**
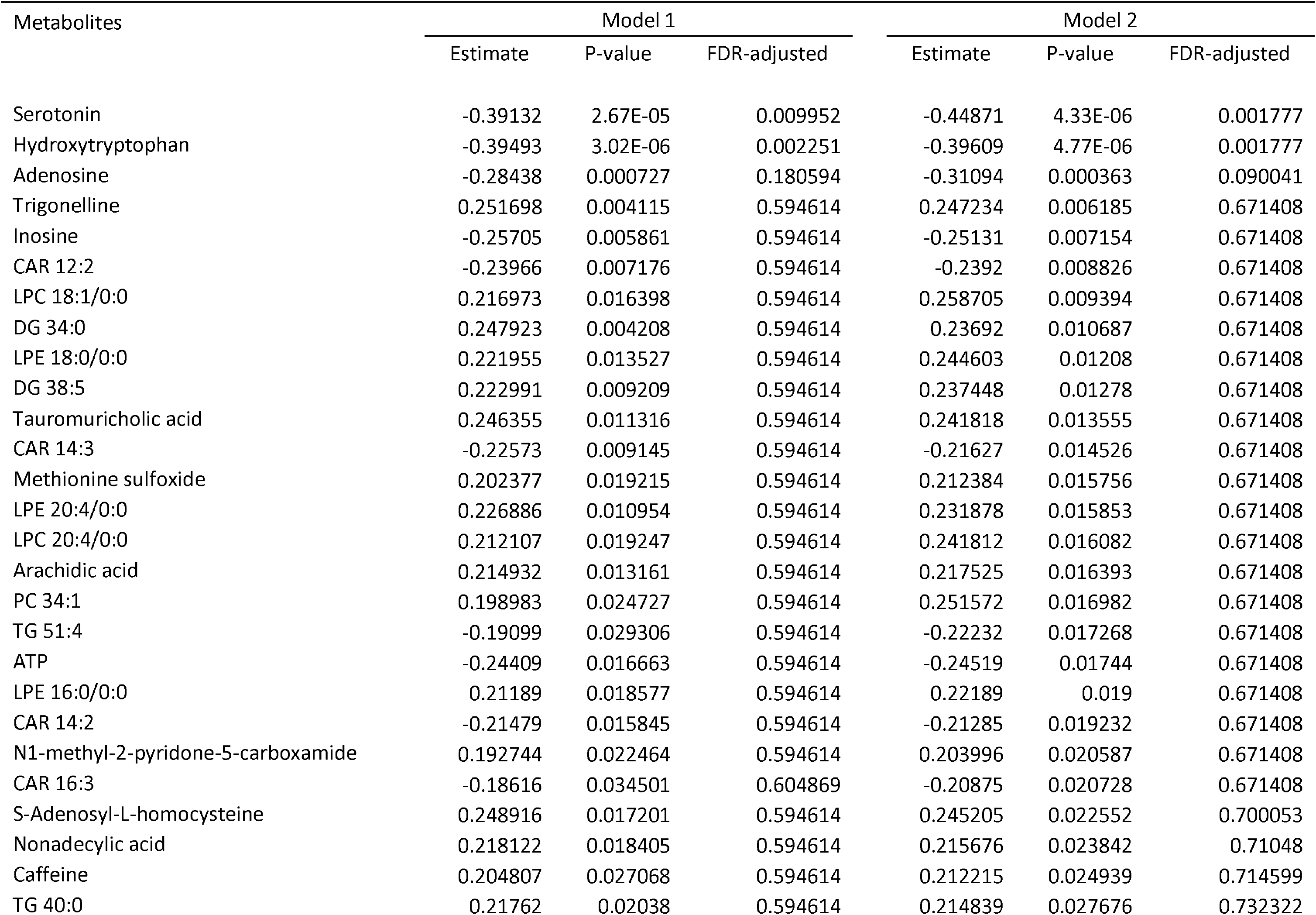
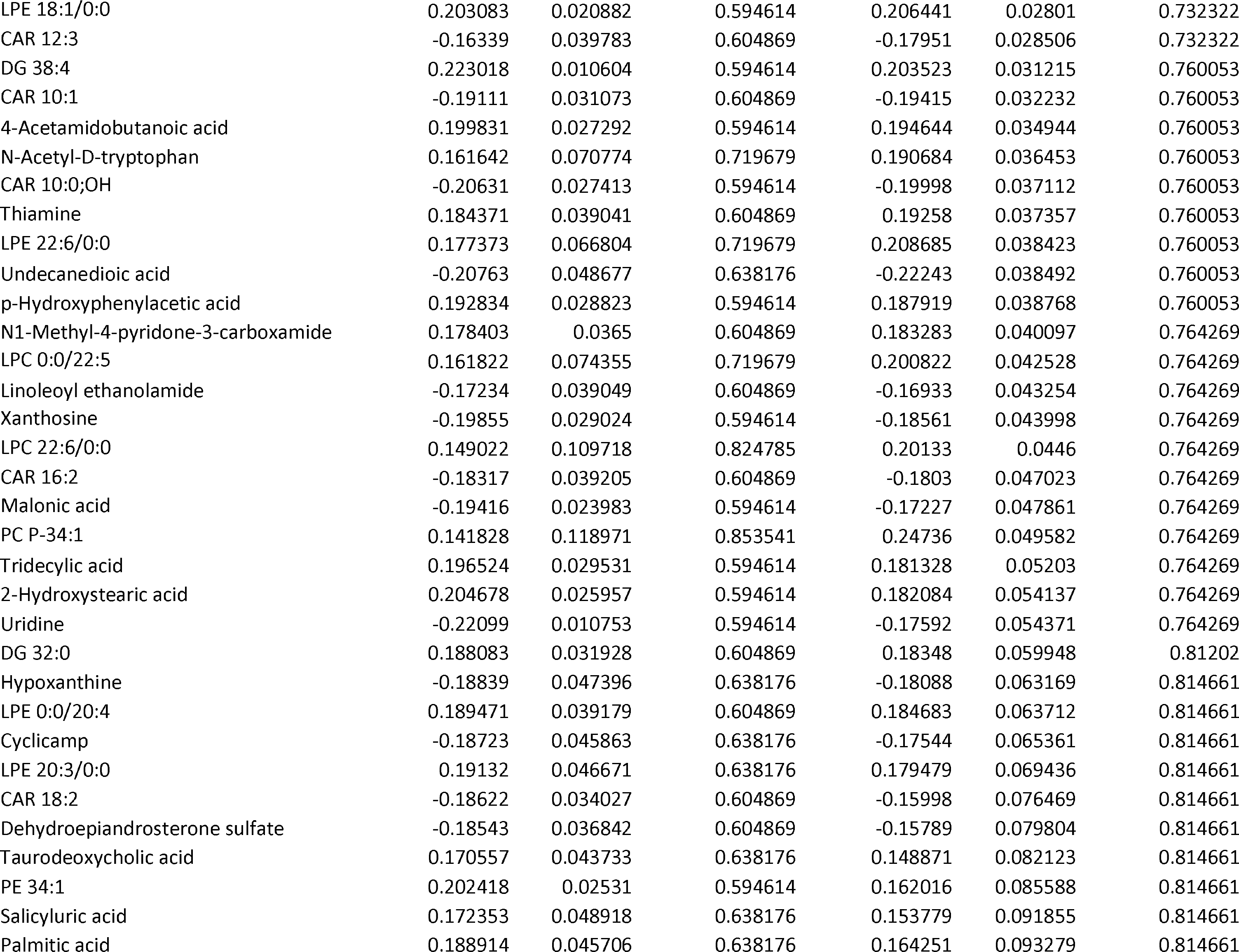
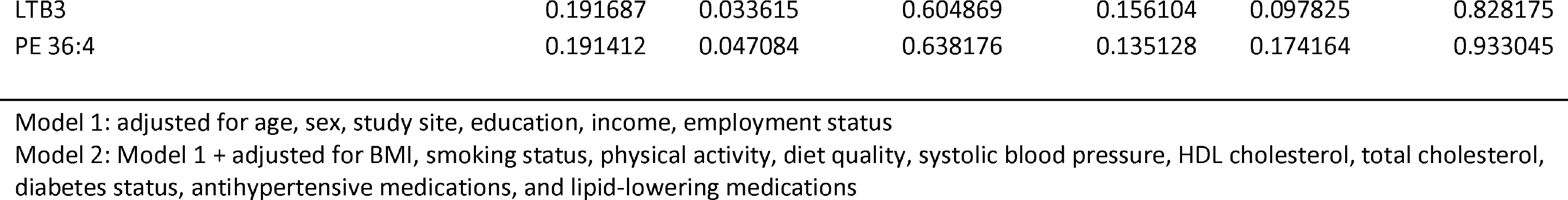
Metabolites nominally associated with depression status among MESA Black participants in either of the two multivariable models.

